# Ultra-Deep Sequencing Reveals the Mutational Landscape of Classical Hodgkin Lymphoma

**DOI:** 10.1101/2021.06.25.21258374

**Authors:** Felicia Gomez, Matthew Mosior, Joshua McMichael, Zachary L. Skidmore, Eric J. Duncavage, Christopher A. Miller, Haley J. Abel, Yi-Shan Li, Kilannin Krysiak, David A. Russler-Germain, Marcus P. Watkins, Cody Ramirez, Alina Schmidt, Fernanda Martins Rodrigues, Lee Trani, Ajay Khanna, Julia A. Wagner, Robert S. Fulton, Catrina Fronick, Michelle O’Laughlin, Timothy Schappe, Amanda Cashen, Neha Mehta-Shah, Brad S. Kahl, Jason Walker, Nancy L. Bartlett, Malachi Griffith, Todd A. Fehniger, Obi L. Griffith

## Abstract

The malignant Hodgkin and Reed Sternberg (HRS) cells of classical Hodgkin lymphoma (cHL) are scarce in affected lymph nodes, creating a challenge to detect driver somatic mutations. As an alternative to cell purification techniques, we hypothesized that ultra-deep exome sequencing would allow genomic study of HRS cells, thereby streamlining analysis and avoiding technical pitfalls. To test this, 31 cHL tumor/normal pairs were exome sequenced to ∼1000x median depth of coverage. An orthogonal error-corrected sequencing approach verified >95% of the discovered mutations. We identified mutations in genes novel to cHL including: *CDH5* and *PCDH7;* novel mutations in *IL4R*, and a novel pattern of recurrent mutations in pathways regulating Hippo signaling. This study provides proof-of-principle that ultra-deep exome sequencing can be utilized to identify recurrent mutations in HRS cells, allowing for the analysis for clinically relevant genomic variants in large cohorts of cHL patients.

## Introduction

Classical Hodgkin lymphoma (cHL) accounts for about 10% of newly diagnosed lymphoma cases and is among the most common cancers diagnosed in adolescents.^1^ While most cHL patients respond to front-line therapy and are cured, a subset of patients relapse or are refractory, and remain a clinical challenge. Moreover, standard treatments, including chemotherapy and radiation therapy, may have serious long-term complications. Although brentuximab vedotin and immune checkpoint blockade have improved outcomes in relapsed/refractory cHL,^2–4^ improved prognostication and targeted treatment options continue to be an unmet need for this malignancy.

In the last decade, high throughput genomic sequencing has provided insight into cancer pathogenesis and has facilitated the development of novel therapies. Collaborative organizations such as the International Cancer Genome Consortium (ICGC) and The Cancer Genome Atlas (TCGA) have sequenced thousands of tumor genomes.^5^ However, cHL was not studied in these efforts. A key barrier to the genomic characterization of cHL is the paucity of the malignant Hodgkin and Reed Sternberg (HRS) cells, which generally comprise <5% of the tumor and are surrounded by an immunosuppressive non-neoplastic immune cell infiltrate.^6^ Only a few studies have investigated the genomic alterations representative of cHL. These studies have addressed the challenges of rare HRS cells in several ways, including the use of HRS-derived immortalized cell lines,^7^ HRS cell isolation from primary samples using laser capture microdissection or flow cytometric sorting,^8–10^ or through the study of circulating tumor DNA.^11,12^ From these studies, patterns of recurrent somatic mutations have been identified within the NF-κβ (e.g., *TNFAIP3*), JAK/STAT (e.g., *STAT6* and *SOCS1*), and the PI3K/AKT (e.g., *ITPKB* and *GNA13*) signaling pathways.^9,11, 13–17^ Mutations that affect *B2M* and impact HLA class I expression are also recurrent in cHL.^7,9,18^ Despite this growing body of work that has provided a preliminary characterization of the genomic landscape of cHL, experimental limitations include the use of cell lines, small sample sets, and biases introduced via complex isolation techniques that require high HRS cell content. Since these techniques will not be feasible in a routine pathology laboratory, alternative approaches that utilize bulk lymph node tissue are required to perform large studies to correlate the impact of somatic mutations on clinical outcomes. Furthermore, the pathologic confirmation of cHL (especially in the relapsed setting) based on tissue from a core needle biopsy remains challenging.^19^ The integration of genomic testing could enhance the diagnostic utility of a needle biopsy, compared to the substantially more invasive excisional lymph node biopsy.

To address these barriers in the field, we performed ultra-deep exome sequencing to discover recurrent genomic events in 31 cHL lymph node biopsies. An approach using multiple independent sequencing libraries per sample and custom variant filtering was developed to overcome the challenge of uncovering recurrent mutations among high-coverage, low variant allele frequency (VAF) sequencing data.^20^ Mutations were validated using an orthogonal error-corrected sequencing technique. This application of ultra-deep exome sequencing to a rare malignant cell population created a reliable and reproducible landscape of cHL somatic mutations, expanding our understanding of the genomic drivers and pathways important in cHL pathogenesis.

## Results

### Patient characteristics

Ultra-deep exome sequencing was used to identify novel recurrent mutations in 31 fresh-frozen cHL biopsies with matched non-malignant tissue. This cohort included samples from 27 (87%) newly diagnosed and 4 (13%) relapsed cHL patients with clinical characteristics shown in Table 1.

**Table 1.** Patient Characteristics and Demographics. Data are presented as No. (%) unless otherwise indicated. Histologic subtype *other* includes one patient where Hodgkin lymphoma (HL) and chronic lymphocytic leukemia were present (CLL; The CLL sample was excluded from all analyses) and a second patient that was designated as interfollicular. Histologic subtype *not specified* indicates that a cHL subtype was not provided. *indicates five samples with no information regarding the number of RS cells.

| **Characteristic** | **Value (N = 31)** |
| --- | --- |
| Female sex | 16 (52) |
| Age, years, median (range) | 36.5 (18 – 69) |
| Histologic Subtype |  |
| Nodular sclerosis | 21 (68) |
| Mixed cellularity | 1 (3) |
| Other | 2 (6) |
| Not specified | 7 (23) |
| Stage |  |
| I | 0 (0) |
| II | 14 (45) |
| III | 7 (23) |
| IV | 8 (26) |
| Unknown | 2 (6) |
| Early Stage Prognostic Group |  |
| Favorable | 2 (7) |
| Unfavorable | 11 (35) |
| Unknown | 3 (10) |
| Not applicable (Stage III/ IV) | 15 (48) |
| Bulky Disease |  |
| No | 21 (68) |
| Yes | 9 (29) |
| Unknown | 1 (3) |
| EBV Status by EBER Stain |  |
| No | 21 (68) |
| Yes | 5 (16) |
| Unknown | 5 (16) |
| Approximate total number of RS Cells*<br>median (range) | 1250 (22 – 6000) |

### Genomic Data Generated and Coverage Statistics

We generated ∼1000x coverage exomes for both the tumor and normal samples for all patients, sequencing three independent libraries for each sample (Methods). The median depth of coverage was 939x (range: 526-1,294x) for normal samples and 1025x (575-1,321x) for tumor samples (Fig. 1a; Supplementary Figure 1). On average 111.11 gb were generated per sample with an average on-target duplication rate of 16.5%, with 92% and 94% of bases covered at 200x, and 62% and 67% of the bases covered at 400x in the normal and tumor, respectively.

**Figure 1.**
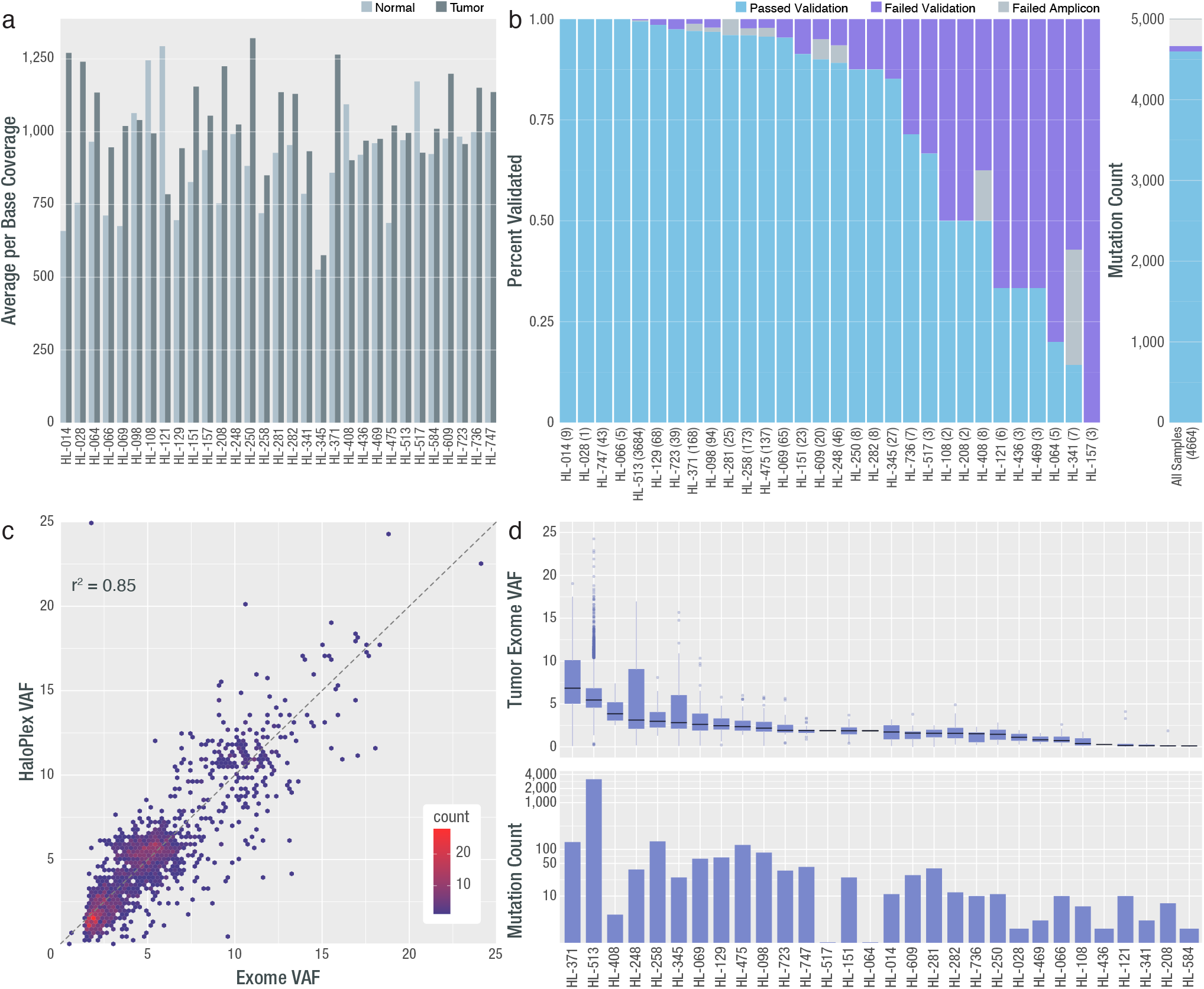
Deep Exome Coverage, Validation Rate, Final Variant Count and VAF per Sample. **a)** Average per base coverage for targeted regions across each tumor and normal sample. b) A targeted orthogonal sequencing strategy (HaloPlex; Agilent) was used to validate all variants that passed filtering strategies for all samples (except for the hypermutated sample, for which a subset of variants were selected for validation-Methods). The percent of assayed variants that passed or failed are shown by sample as well as the number for which validation was not possible due to amplicon design failure. The overall validation rate by mutation count is also shown at the right. Note: One patient (HL-584) did not have any variants pass filtering and review, therefore 30/31 patients were included in validation. c) Comparison of exome VAF and HaloPlex VAF is shown for samples with 2 or more variants. d) VAF and variant count for all variants across all samples used in all further analyses. Note: Variants from one patient failed validation (HL-157) and no further variants were called in de novo exercises. This patient was removed from all further analyses. Additionally, the patient who did not have variants to validate, gained 2 variants in the de novo exercise and was included in the final cohort (HL-584).

### Initial Variant Analysis and Variant Validation

After variant calling, filtering, and manual review, we identified 4,692 SNVs and INDELs from the ultra-deep exomes that were considered for validation. We identified one relapse sample (HL-513) with 3,684 mutations (78% of all variants from the cohort), suggestive of a hypermutator phenotype.^21,22^ Excluding this hypermutated patient, the median number of somatic mutations across the cohort was 11 and the mean was 32 (range 0-148). The VAFs for all sites identified, including variants from the hypermutated patient, were consistent with detection from rare HRS cells (mean VAF=5.7%; median VAF=5.2%; range=0.5-24.1%).

Following the identification of variants from the ultra-deep exomes, a HaloPlex panel, with molecular barcodes (Unique Molecular Indexes, UMIs) included for error-correction, was designed to validate all variants from all patients, except a subset of sites from the hypermutated patient. Due to HaloPlex panel size limitations, a subset (834/3684 randomly selected) of the hypermutated patient mutations were included. A total of 19,392 amplicons were generated, with these probes spread over 1842 SNVs and INDELs, as well as 327 exons of 30 genes (Supplementary table 5). High depth sequencing data was generated for HaloPlex targeted regions, with a median of 224,355,855 total reads/sample (range 125,968,046-387,141,061 total reads) and a median error-corrected depth of coverage at 2,168x and 3,971x in the normals and tumors, respectively.

The overall exome call validation rate by HaloPlex was 96.7% (1754/1814) (Fig. 1b; Supplementary Figure 2). This includes 1,405 fully validated sites and 349 “tumor only” validated (where matched normal data were unavailable -Methods). This rate also accounts for sites that could not be evaluated due to low tumor read depth (1.5%; 27/1842 sites) or HaloPlex amplicon failure (0.05%; 1/1842). An additional 2,850 sites from the hypermutated patient were not assayed due to HaloPlex size limitations (Methods). These sites were included in subsequent assays because 811/823 (98.5%) of tested HL-513 mutations were validated (this rate accounts for sites with low tumor depth; 11/834). A total of 60 sites (3.3%; 60/1814) failed validation. The correlation of the HaloPlex and exome VAFs among validated sites in samples with more than 2 variants was R^2^=0.85 (p<0.01), demonstrating significant concordance of VAF between the two methods (Fig. 1c).

After validating exome-discovered mutations using HaloPlex, new variants were discovered across our entire HaloPlex target space (Supplementary Table 5). This included 10 known cHL hotspots (Methods; Supplementary Table 1). From this *de novo* variant calling, 135 new sites were identified across the HaloPlex target region including 7 new variants at known HL hotspots. After updating the annotations of all potential variants (*de novo* and exome-validated) (Methods) the final annotated dataset consists of 4,116 non-synonymous coding somatic mutations that were carried forward for all subsequent analyses (Fig. 1d, Supplementary Table 6). The final cohort includes 30 individuals, as one patient, of the original 31, had 3 exome-discovered variants that all failed validation (patient HL-157). The median number of sites in this final validated data set was 11, and the mean was 32.9 (range: 1-148), excluding the hypermutated patient, who contributed 3,160 variants to the final data set. The mean and median VAFs were 5.6 and 5.1, respectively (range: 0.03-24.10) (Fig. 1d). Among the relapse samples, excluding the hypermutated sample, we observed a median of 10 variants and a mean of 19 variants (range inclusive of hypermutated sample: 10-3160). There were 27 genes with variants in at least one relapse sample (not including genes only mutated in the hypermutated sample; Supplementary Figure 3).

### Recurrent and Significantly Mutated Genes

A total of 3,168 somatically mutated genes were identified across all 30 samples, versus 732 mutated genes when the hypermutated patient was excluded. Somatic mutations were identified in 263 genes in at least 2/30 samples. The most recurrently mutated genes in our cohort are *SOCS1* [43.3%], *TNFAIP3* [40%], and *IGLL5* [26.7%] (Figure 2). We identified 28 genes that were significantly mutated above the background mutation rate (significantly mutated genes; SMGs; Fig. 2). SMGs identified in this analysis included several genes that have previously been shown to be mutated in cHL, including members of the JAK/STAT signaling pathway (i.e. *SOCS1* and *STAT6* [20%]) and the NFκB signaling pathway (i.e. *TNFAIP3* [40%] and *XPO1* [20%]). Other SMGs identified in this study known to be mutated in cHL include: *B2M* [16.7%], *ITPKB* [16.6%], and *GNA13* [20%]. Components of the SWItch/Sucrose Non-Fermenting (SWI/SNF) complex including *BCL7A* [13%], *SMAD3* [6.6%], and *ARID1A* [6.6%], were also mutated. *BCL7A* and *SMAD3* are known to be involved in lymphomagenesis but not previously implicated in cHL. *ARID1A* was not among our SMGs, but was mutated in 3 cases and has been identified by others to be recurrently mutated in cHL.^9^ A summary of genes found to be recurrently mutated in cHL across several recent studies of adult cHL^9–11,18^ is provided (Supplementary Figure 4). To our knowledge, the following SMGs have not previously been reported in cHL: *AXDND1* [6.7%], *CDH5* [13.3%], *LIMD2* [10%], *OR13C2* [6.7%], *PCDH7* [20%], *RDH12* [6.7%], *SCN9A* [6.7%], and *STRAP* [6.7%].

**Figure 2.**
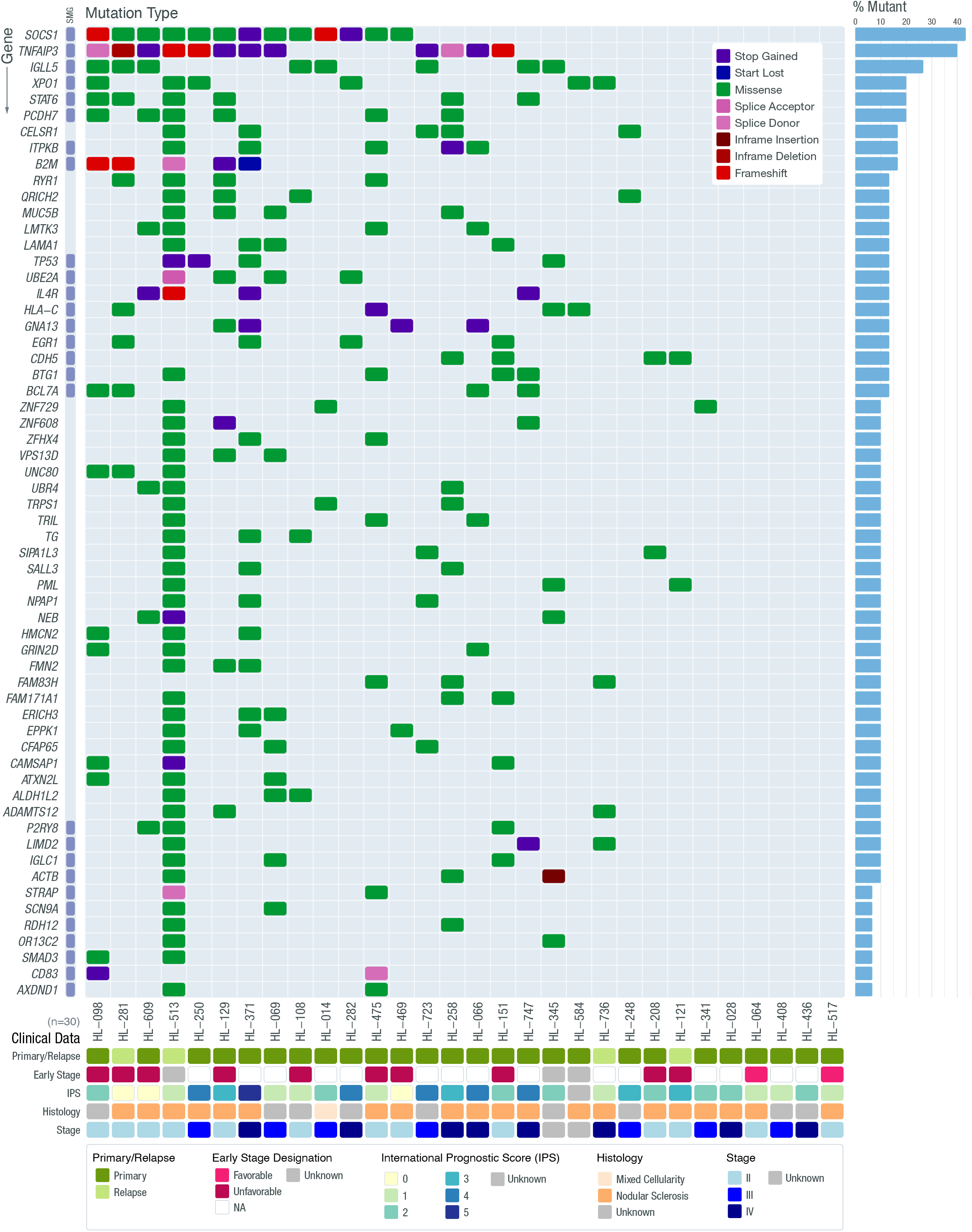
Recurrently Mutated Genes in Hodgkin Lymphoma. The frequency and type of mutations affecting genes mutated in 3 or more patients in our cohort are shown in each row (60 in total). Genes determined to be significantly mutated using MuSiC (FDR < 0.05, minimum of convolution and likelihood ratio tests) are highlighted (SMG = Significantly Mutated Gene). Each column represents a patient in the cohort. The bar graph on the right summarizes the frequency of mutations for that gene across the entire cohort. For genes with multiple mutations in a single patient, only 1 mutation type is shown prioritizing the most severe mutation (listed in order in the legend).

**Figure 3.**
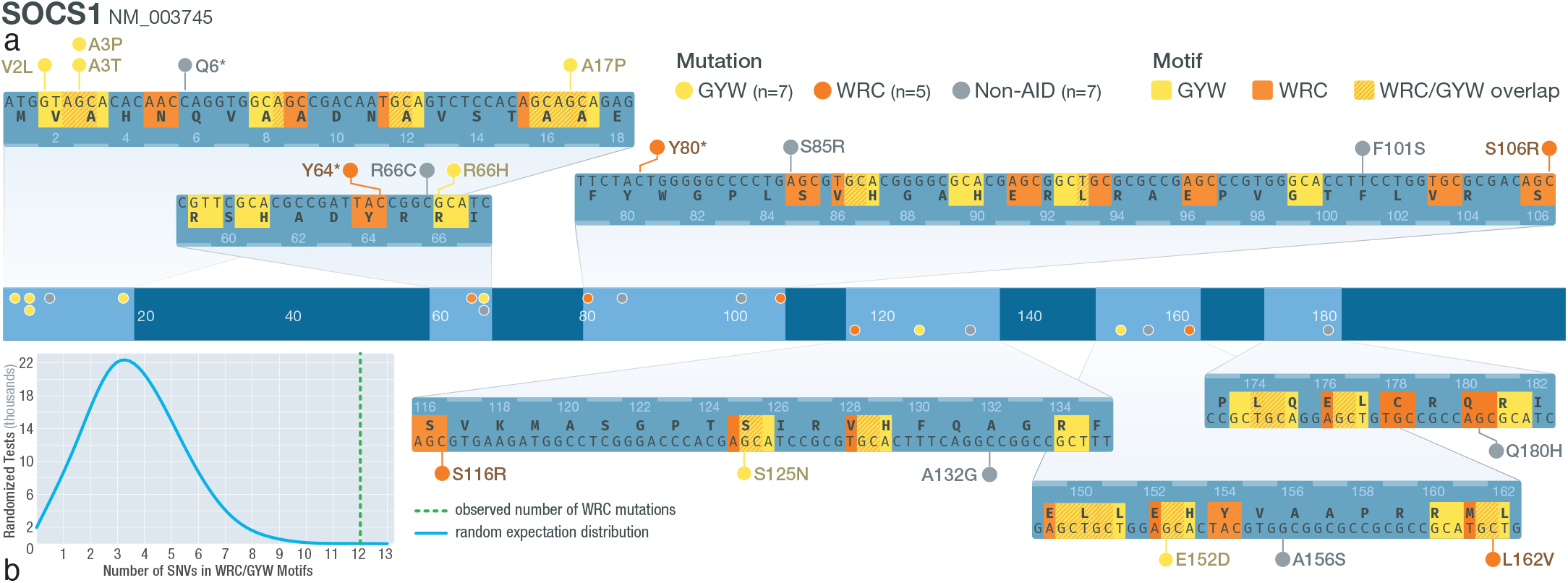
Mutations in AID motifs at *SOCS1*. a) Schematic of *SOCS1* gene structure. Middle track shows where *SOCS1* SNVs are located. Zoomed-in tracks (top and bottom) show the location of WRC/GYW motifs and the mutations that lie within or outside motifs (W= A/T; R= A/G; Y= C/T). b) Distribution of counts of simulated mutations observed at SOCS1 that were in WRC motifs. The actual observed number of WRC SNVs (12/19) is shown using a green dashed line.

Previously unreported mutations in multiple cadherin genes were identified. There were 74 mutations in 42 different cadherin genes identified. The two cadherin SMGs were *PCDH7*, a protocadherin, and *CDH5*, a type II classical cadherin. Many of the cadherin mutations were only identified in the hypermutator patient; however, if these are excluded, 21 mutations were discovered across 9 different cadherin-related genes: *CDH10* [3%], *CDH23* [3%], *CDH5* [16%], *CELSR1* [13%], *DSC2* [3%], *DSG3* [7%], *FAT3* [3%], *PCDH19* [3%], and *PCDH7* [16%].

### Mutation Signature Analysis and AID Targets

To further understand the etiology of somatic mutations in cHL, a mutation signature analysis was conducted to identify the trinucleotide context in which the somatic mutations occurred. The patterns of SNV mutational classes identified were compared to the COSMIC v2 database of known mutational profiles.^23^ The most prevalent mutation signatures were 1, 3, 9, and 30. Additionally, signature 6 was observed in the hypermutated patient (Supplementary Figure 5).

We observed the presence of COSMIC signature 9 in several patients (Supplementary Figure 5). This signature is associated with the activity of an enzyme called <u>a</u>ctivation-induced cytidine <u>d</u>eaminase (AID). In a recent analysis of the CLL mutational landscape, two AID signatures were observed;^24^ one that is characterized by a canonical AID signature that includes C to T/G mutations at the WR<u>C/G</u>YW AID hotspot, as well as a non-canonical AID signature that includes A to C mutations at WA motifs, which is consistent with COSMIC signature 9. Although our analysis suggests that signature 9 (non-canonical AID) is common in our cohort, the COSMIC v2 database does not contain an AID signature that is associated with canonical AID activity. As shown by Kasar et al.,^24^ mutation signatures associated with canonical and non-canonical AID activity share some similarities; thus it is possible that our identification of signature 9 could in fact reflect the presence of canonical AID activity. To address whether our data actually indicate a canonical AID signature, a list of genes known to be significant targets of off-target canonical AID activity in diffuse large B-cell lymphoma (DLBCL) and follicular lymphoma (FL) was generated.^25,26^ Our analysis revealed 24 canonical AID-target genes were mutated in at least one non-hypermutated patient sample. Of these, 8 genes had SNVs (23 SNVs total) located in the known WR<u>C/G</u>YW AID target motif, including: *SOCS1*, *IGLL5, ARID5B, CD83, HIST1H2AL, ZFP36L1, HIST1H1B, and HIST1H1C*. Of the 19 SNVs identified in *SOCS1*, 12 were within WR<u>C/G</u>YW motifs. Four of 9 SNVs in *IGLL5* and two of two SNVs in *HIST1H1B* were within a WR<u>C/G</u>YW motif. At *ARID5B, ZFP36L1, HIST1H2AL,* and *HIST1H1C*, 1 SNV was identified at each locus in a WR<u>C/G</u>YW motif (1/1; 1/3; 1/1; 1/1). To test whether the number of mutations identified within WR<u>C/G</u>YW motifs was significantly different from random expectation, we simulated the number of mutations we observed at each gene 100,000 times and asked how many times mutations were identified in a WR<u>C/G</u>YW motif. On average, each *SOCS1* simulation had 3.73 mutations that met the criteria for a potential mutation generated by aberrant AID activity. In contrast, only 9 simulations had at least 12 variants, as we observed in our actual cohort (p=0.0009). Following a similar logic, the simulated *IGLL5* dataset identified 47/100,000 tests with at least 4 SNVs that could result from aberrant AID activity (p=0.0005). We assessed the enrichment of off-target AID mutations at the other loci we identified and three additional genes that exhibited significant enrichment for AID mutations: *ARID5B* (p=0.0083), *HIST1H1B* (p=0.017) and *ZFP36L1* (p=0.014). We also asked whether the overall number of mutations we saw across the 24 potential AID targets was different from random expectations. The results of this permutation analysis showed that across the coding space of all 24 genes, we would expect to observe 6.36 total mutations meeting our AID target criteria. We did not observe any permutations with a total of 23 mutations (p<0.00001), suggesting that the overall pattern of off-target AID activity is significantly different from random expectations. These results indicate that we observed a significant number of mutations that are the result of canonical off-target AID, across multiple loci and specifically at *SOCS1* and *IGLL5*.

### JAK/STAT Signaling Mutations

Mutations were identified in 22 genes from the JAK/STAT signaling pathway, with 50% of our cohort (15/30) having at least one somatic mutation in a JAK/STAT signaling gene. Several novel stop gain and frameshift mutations were discovered, clustered in the genomic region that encodes the cytoplasmic region of IL-4R that contains an immunoreceptor tyrosine-based inhibitory motif (ITIM). These mutations were downstream of the box1 motif (JAK1 interaction region) and the amino acids that are thought to be required for STAT6 interaction.^27^ The potential loss of function mutations we have identified may represent an additional mechanism to promote cHL proliferation in response to IL-4 stimulation via mutation of normally inhibitory ITIMs.

Two nonsense mutations were also identified in the cytoplasmic regions of CSF2RB (IL3RB). This gene encodes the common β chain (CD131) that associates with the IL-3, IL-5, and the granulocyte-macrophage colony stimulating factor (GM-CSF) alpha receptors. This gene has been shown to be recurrently mutated in HL cell lines^7^ and HL primary samples.^9,28^ The nonsense mutations we identified at this locus are beyond the JAK2 box 1 motif (amino acids 474-482). Because a truncated isoform of the common β chain may be related to the pathogenesis of acute myeloid leukemia (AML),^29^ and mutations in the cytoplasmic region have been associated with growth in T-cell acute lymphoblastic leukemia,^30^ it is possible that these mutations are related to cHL pathogenesis.

### Regulating Hippo

The Hippo pathway is highly conserved across many species, playing a role in cell death, differentiation, and inhibition of cell proliferation.^31^ Somatic mutations in 31 genes involved in pathways regulating Hippo/TAZ/YAP or directly interacting with the Hippo cascade were identified. These genes are mutated in 40% (12/30) of our cohort. Two SMGs, *CDH5* and *GNA13*, are among this group. CHD5/VE-cadherin has not been previously described as a driver of cHL. VE-cadherin is linked through its cytoplasmic tail to adherens junction (AJ) proteins, p120, Beta-catenin, and plakoglobin.^32^ The mutations identified here span amino acids 650 - 680, which could impact the p120 and Beta-arrestin association regions of VE-cadherin that are vital for the stability of catenin-cadherin complexes (Fig. 4). It has been shown that disruption of VE-cadherin clustering or suppression of VE-cadherin expression results in the nuclear localization of YAP and the promotion of cell proliferation.^33,34^

**Figure 4.**
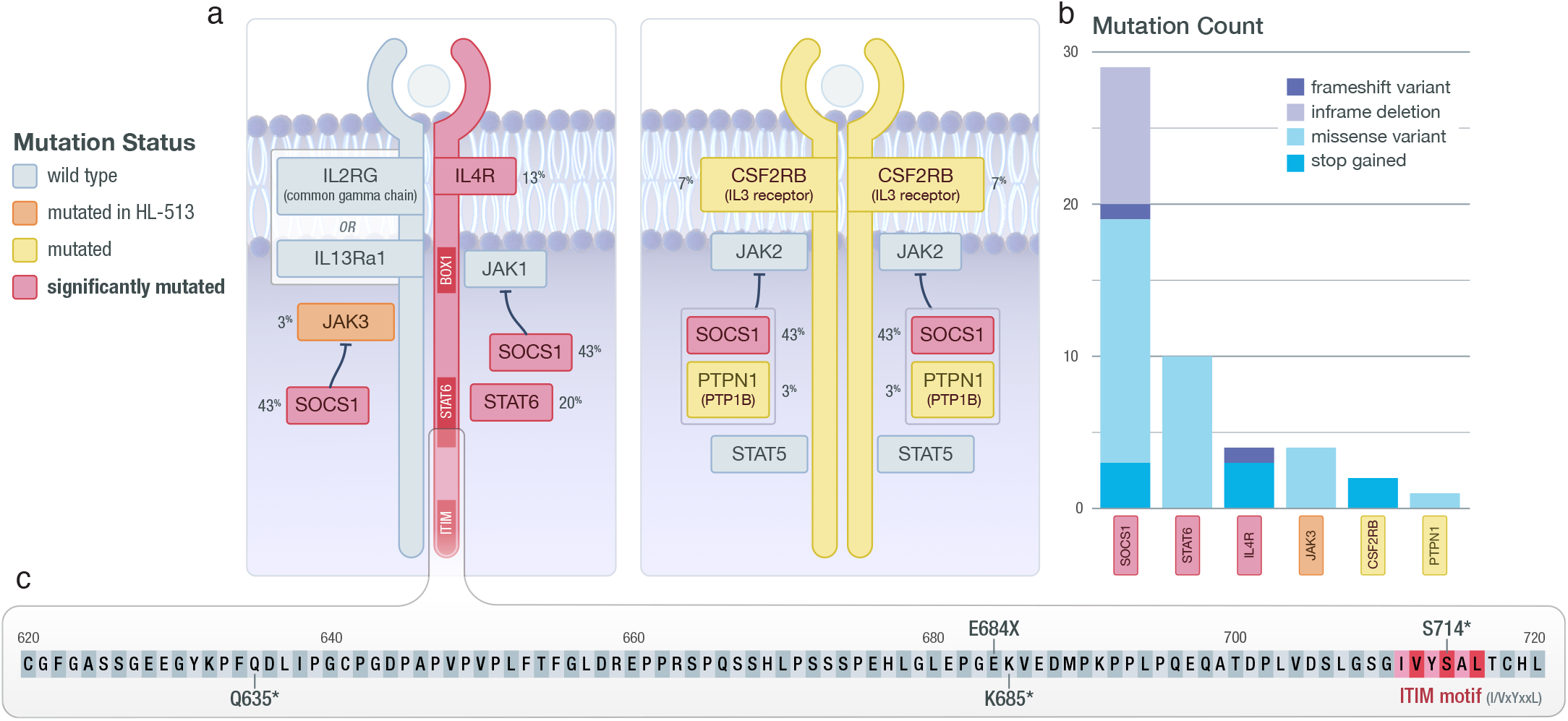
JAK/STAT Signaling. a) Diagram of components of the JAK/STAT signaling cascade. Identified SMGs are shown in red; genes mutated in at least one non-hypermutated sample are shown in yellow; genes mutated only in the hypermutated sample (HL-513) are shown in orange. The gene mutation frequency across the cohort is shown as a percent. b) The total number and type of mutation observed are shown. c) Zoomed-in view of the C-terminal region of IL4R, where the observed truncating mutations are located. Also shown is the proximity of identified mutations to an ITIM motif that may impact STAT6 activation.

**Figure 5.**
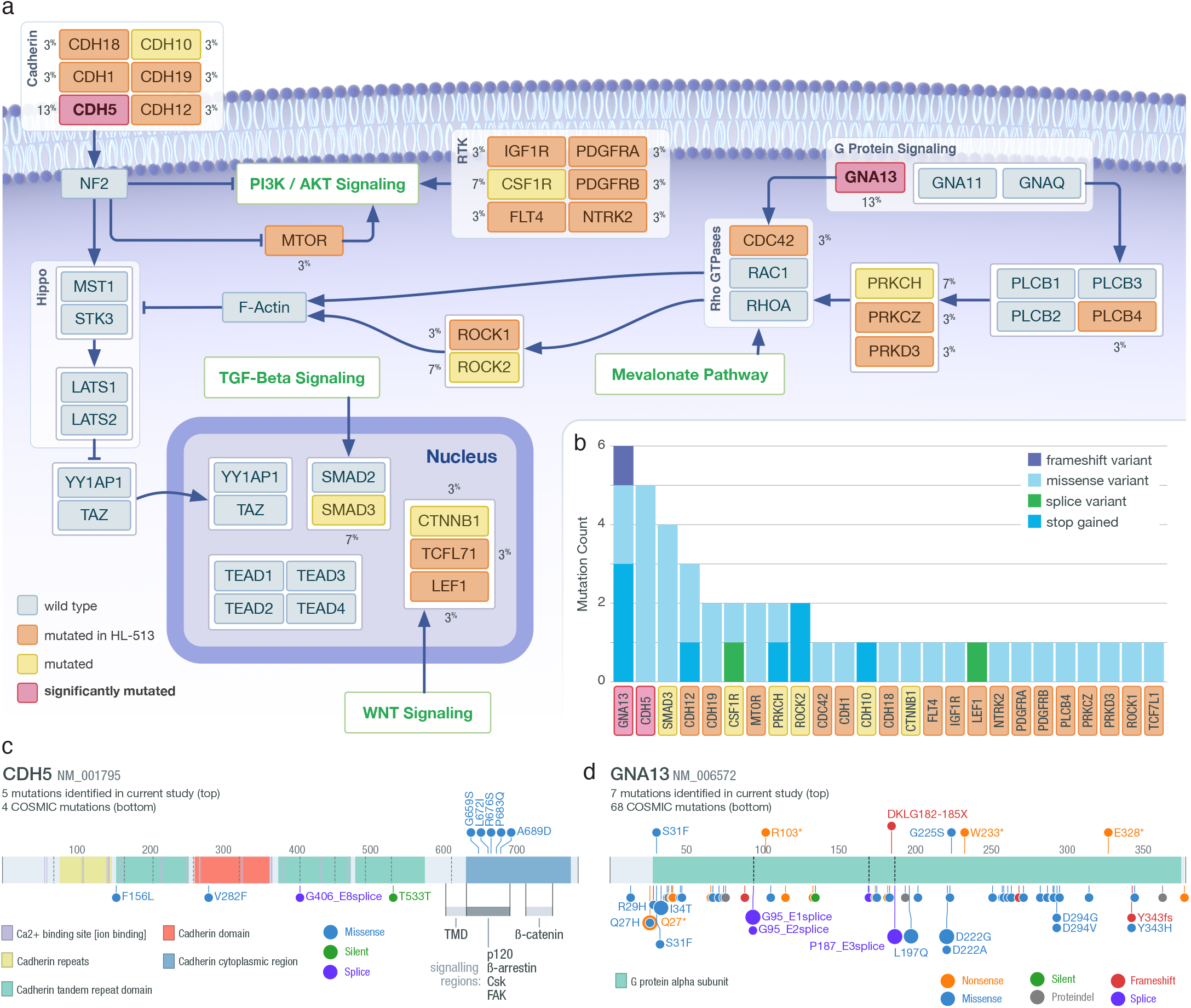
Regulating Hippo. a) Diagram of pathways that regulate Hippo signaling. Identified SMGs are shown in red; genes mutated in at least one non-hypermutated sample are shown in yellow; genes only mutated in the hypermutated sample (HL-513) are shown in orange. The gene mutation frequency across the cohort is shown as a percent. The pathways labeled in green indicate larger pathways not shown in this diagram. b) The total number and type of mutation observed are shown in the inset barchart. c) Lolliplot of *CDH5*; mutations identified in the current study are shown on the top and COSMIC mutations found in lymphoid tissue on the bottom. d) Lolliplot of mutations identified at *GNA13*; mutations identified in the current study are shown on the top and COSMIC mutations found in lymphoid tissue on the bottom

Gα13 (encoded by *GNA13*) is a g-protein coupled receptor known to be mutated in cHL.^9–11,18^ Gα13 is also involved in G-coupled signaling that activates Rho GTPases, which subsequently activates Rho-associated protein kinase I and II (ROCK1/2). This leads to actin cytoskeletal tension, and has been shown to negatively regulate YAP/TAZ phosphorylation.^35–38^ We observed several missense, frameshift, and nonsense mutations, consistent with previous observations in cHL,^9–11^ Burkitt’s lymphoma, and DLBCL.^39,40^ Frameshift and nonsense mutations may cause loss of function of Gα13, which is unlikely to promote TAZ/YAP signaling.^39,41^ However, two missense mutations were observed, including one located at G225S, which is similar to a dominant-negative mutation at G225A^40,42^ and close to Q226L, which is known to cause constitutive activation of Gα13.^36,43,44^ In addition to the likely loss of function consequences of the *GNA13* mutations observed, we identified other variants that may impact Hippo signaling either through GTPases or other pathways, including mutations at *PRKCH*, *ROCK2,* and *CSF1R*.^45–47^

### MAPK Signaling Mutations

We identified mutations in 59 genes that are involved in MAPK signaling pathways. Moreover, 43% (13/30) of the cohort had at least one mutation in a gene annotated to a MAPK signaling pathway (Supplementary Figure 6). Mutations in *DUSP4* (missense mutation) and *DUSP6* (single base deletion) were also defined. DUSP6 and DUSP4 are known to inhibit (dephosphorylate) ERK.^48–50^ If the mutations we identified alter the function of DUSP6 and DUSP4, they would support the proliferation of HRS cells. Mutations in the MAPK3 kinases that precede p38 in the signaling cascade (*MEK3*, *TAOK1/2*, and *MAP3K6*) and in scaffold proteins like *KSR1/2* and *JIP1/2* were discovered. These results suggest that previously unreported key growth pathways are mutated in cHL and may contribute to the pathogenesis of cHL.

### Phosphatidylinositol Signaling Mutations

The PI3-kinase (PI3K)-Akt signaling pathway is an important regulator of many cellular functions and is often targeted and dysregulated in cHL.^17,51,52^ Thirty percent (9/30) of our cHL patient cohort had a mutation in a gene mapped to phosphatidylinositol signaling, including the SMG *ITPKB*. Similar to Tiacci et al,^10^ *ITPKB* was mutated in 16.6% of our cohort (Supplementary Figure 7). Several missense mutations and one nonsense mutation (p.Y4*) were discovered, which is in contrast to the high frequency of truncating mutations previously reported.^10^ We also identified several mutations in this pathway that are involved in calcium signaling (*ITPR1*, *ITPR3*, and *CALM2*). Although these are not well described drivers of cHL, it has been shown that these genes can impact oncogene-induced senescence and can promote cellular proliferation.^53^

### Germline Mutation in Hypermutated Patient

To address the etiology of the hypermutated patient, the patient’s germline was analyzed for predisposing genetic conditions that could explain the hypermutated phenotype. Since a strong presence of COSMIC mutation signature 6 was observed, which is generally associated with defects in DNA mismatch repair (MMR) and microsatellite instability (MSI), analysis for unique mutations in mismatch and base excision repair genes was performed (Methods and Supplementary Table 2). Germline mutations in *NTHL1* and *MSH6* were discovered. The *NTHL1* mutation is an SNV in exon 1 causing a stop gain at Q287*. The *MSH6* mutation identified was a 58 base pair duplication in exon 9, resulting in a frameshift (NM_000179; NP_000170.1:p.Lys1325SerfsTer2). This mutation appears the most likely candidate responsible for the hypermutated phenotype. This duplication is similar in kind and location to a number of the frameshift and nonsense mutations reported in ClinVar that are known to cause Lynch Syndrome, an autosomal dominant cancer predisposition syndrome characterized by MMR deficiency and MSI (Fig. 6)

**Figure 6.**
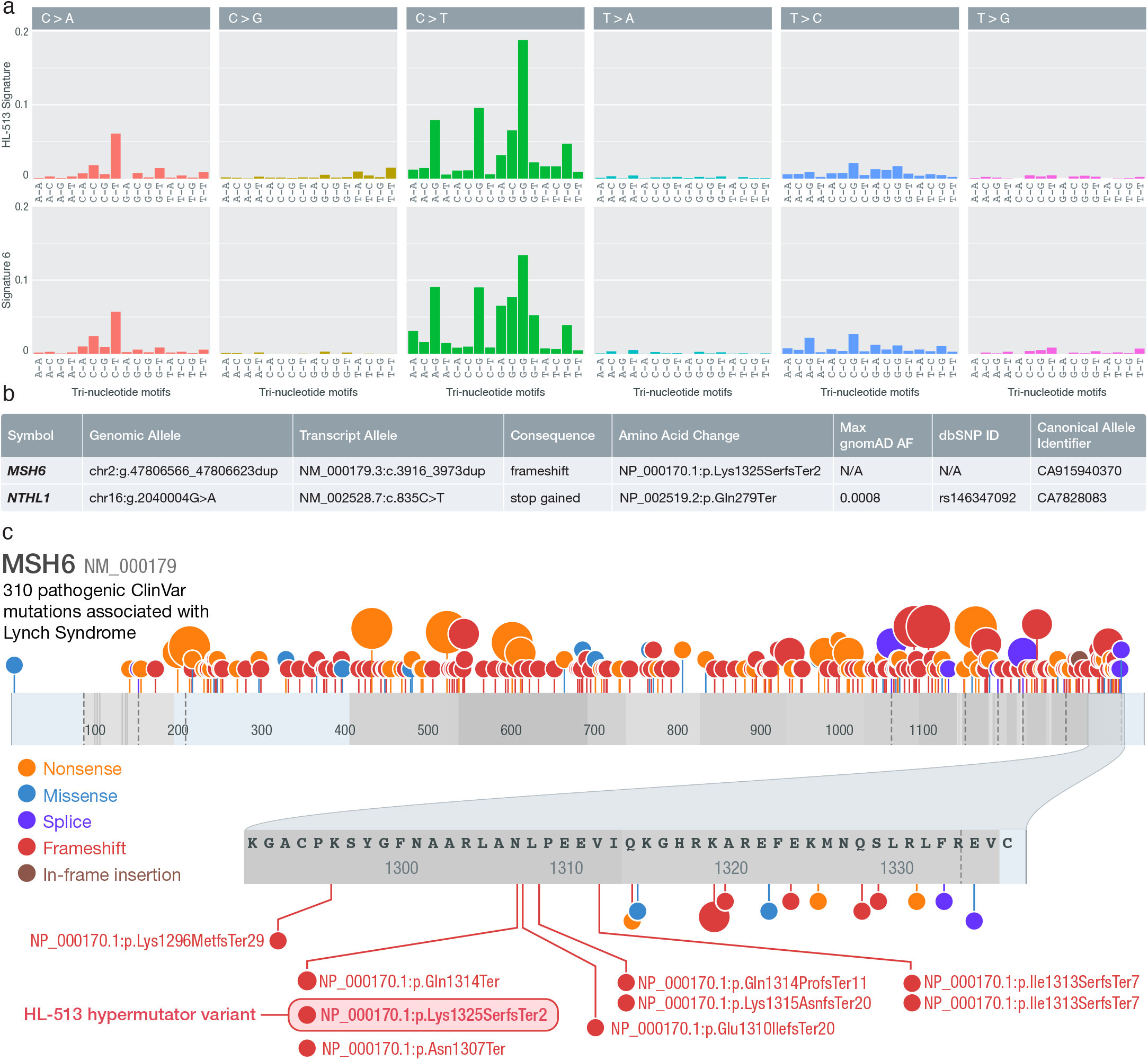
Hypermutated Patient Germline Analysis. a) Comparison of tri-nucleotide sequence contexts of COSMIC signature 6 and HL-513 suggesting a high degree of similarity between the two. b) Summary of the mismatch repair germline mutations identified in HL-513. c) Lolliplot of Lynch Syndrome mutations from ClinVar and the HL-513 mutation identified here. ClinVar mutations annotated as pathogenic and associated with Lynch Syndrome are shown. The shaded oval highlights the duplication identified in HL-513. Note: mutations are plotted by ProteinPaint according to (left-shifted) genomic coordinates but labeled with their HGVS expressions (right-shifted) from ClinVar explaining the discrepancies in some amino acid positions displayed

## Discussion

This study demonstrated the utility of ultra-deep sequencing to uncover recurrently mutated genes in low frequency malignant cells and to further define the landscape of somatic mutations in cHL. We generated ∼1000x exomes in a cohort of 31 primarily newly diagnosed cHLs and matched non-malignant germline tissues. These findings were validated with orthogonal error-corrected sequencing with a recovery rate >95%. Ultra-deep sequencing strategies such as this have been employed to detect low VAF and/or subclonal variants in a number of cancers including ovarian cancer,^54^ CLL,^55^ and AML.^56,57^ However, here we pioneered this approach to overcome the progress-limiting rarity of HRS cells in cHL. These findings open up new avenues of research, as this approach provides a platform for future prospective studies to analyze mutations in bulk cHL biopsies. Ultra-deep sequencing of cHL bulk biopsies confirmed the importance of previously reported pathways in cHL including JAK/STAT, NFkB, and those relevant for immune evasion. Moreover, mutations in previously unreported genes (*AXDND1*, *CDH5*, *LIMD2*, *OR13C2*, *PCDH7*, *RDH12*, *SCN9A*, and *STRAP*) and pathways (Hippo/YAP and MAPK) not often associated with cHL were discovered. Mutations associated with off-target AID activity that may be driving the pathogenesis of cHL were also revealed. Finally, we identified a hypermutated cHL patient and discovered a mutation in *MSH6* that is similar in kind and location to other variants associated with Lynch syndrome, which could be driving the patient’s hypermutated phenotype.

This study identified both known and novel mutations in cHL, with several previously reported JAK/STAT pathway mutations confirmed, including *STAT6*. Truncating mutations in, and proximal to, the ITIM located at the C-terminus of IL-4R were discovered, revealing a novel potential mechanism for constitutive activation of STAT6 in cHL, by eliminating this suppressive function of IL-4R signaling. Kashiwada, et al.^58^ found hyperproliferation in response to IL4 stimulation when the ITIM in IL-4R is disrupted through site-directed mutagenesis in a murine bone marrow cell line, and this response was correlated with increased activation of STAT6. We also observed stop gain mutations in IL3RA. It has been shown that HL cell lines express high levels of IL3RA and also express IL3RB,^59,60^ but there is some dispute as to whether or not HL growth is impacted by the presence of IL3.^59,61^ It has also been suggested that mutations at IL3RB may impact leukemia pathogenesis.^29,30^ Further study is needed to understand whether the mutations we identified support the growth of HRS cells or impact cHL biology.

The most recurrently mutated gene we uncovered is *SOCS1*, a finding observed by others,^9,10^ suggesting concordance of our ultra-deep bulk exome approach with HRS purification approaches. This included a number of frameshift and nonsense mutations, as well as missense mutations, consistent with loss-of-function. SOCS proteins negatively regulate the JAK/STAT pathway, with loss-of-function leading to augmented JAK/STAT growth signaling. Several *SOCS1* mutations were identified in the AID sequence recognition motif, and our analyses suggested off-target AID activity as a driver of some mutations. AID is a crucial enzyme involved in immunoglobulin gene (Ig) somatic hypermutation and class switching that can act on non-Ig chromosomal locations and mutate these regions. Off-target canonical AID activity may be responsible for the pathogenesis of some lymphomas and leukemias.^62–64^ Mottok et al. have implicated aberrant AID activity resulting in off-target somatic hypermutation as the mechanism of SOCS1 mutations in multiple germinal center lymphomas.^65,66^ In addition to AID mutations in *SOCS1* we also show that potentially aberrant AID mutations are present in *ARID5B, IGLL5*, *ZFP36L1,* and *HIST1H1B*. The results of our study suggest aberrant off-target AID activity is a characteristic of cHL and impacts several loci including *SOCS1*, which is the gene most impacted by recurrent somatic mutations in our cohort.

Mutations in 31 genes involved in Hippo regulation were identified, with 40% of our cohort (12/30) having at least one mutation in a Hippo regulating gene. A number of studies have suggested that human tumors use YAP/TAZ, which are integral transcriptional coactivators within the Hippo pathway, to facilitate proliferation, progression, migration, and metastasis.^67^ A recent study demonstrated the importance of YAP/TAZ somatic mutation to squamous cell cancers including: cervical, lung, head and neck, and bladder urothelial squamous cell carcinoma.^68^ High expression of YAP was also shown to be significantly correlated with disease progression and poor prognosis in DLBCL and knock down of YAP expression suppressed cell proliferation in DLBCL cell lines.^69^ Furthermore, the overexpression of MST1 or the knock down of YAP inhibited cell proliferation, promoted cell cycle arrest, and apoptosis in natural killer/T-cell lymphoma.^70^ The most prominent mutated genes we identified that impact Hippo signaling are *CDH5* and *GNA13*, both of which were shown to be SMGs. CDH5, like many other cadherins, is involved in the adherens junction that plays a role in cell architecture and Hippo pathway regulation. We identified a cluster of *CDH5* mutations that are located in amino acids involved in CDH5/VE-cadherin’s interaction with p120, beta-catenin, and plakoglobin. These associations are crucial for the stability of the adherens junction, which if disrupted could facilitate nuclear localization of YAP and TAZ. All of the mutations we identified in *CDH5* are missense, so further work is needed to understand the functional impact of these mutations, but the clustered nature of the mutations suggests that this region is being targeted in cHL. If the mutations identified here impact the stability of the cadherin-catenin complex, they may impact YAP or TAZ nuclear localization and in effect contribute to cell proliferation.

MAPK signaling cascades were also found to be perturbed in the analysis, with 43% of the study cohort having a mutation in a gene annotated to the MAPK pathway. Mutations in MAPK3 and MAPK2, MAPK kinases as well as mutations in scaffold proteins and inhibitors of ERK1/2 were observed. While most of the evidence of MAPK kinase pathway involvement in cHL derives from cell line studies, limited evidence has corroborated this with primary cHL samples.^71–74^ Zheng et al. showed that the phosphorylated form of ERK1/2 is aberrantly active in cultured and primary HRS cells.^74^ They also show that when upstream MAPK kinases (i.e. MEK1/2) are inhibited, the phosphorylation of ERK is inhibited thereby decreasing the proliferation of HRS cells. The results of the current study corroborate the importance of the MAPK kinase pathway in cHL.

A single patient contributed over 70% of the overall somatic mutation burden in this cohort. Since hypermutated phenotypes are often a result of germline predisposition,^21^ we searched for a germline variant that may be responsible for this phenotype. Two mutations were identified, one SNV in *NTHL1* and a 58-base repeated sequence in *MSH6*. Inactivating mutations in *MSH6*, a DNA mismatch repair gene, are associated with Lynch Syndrome. The frameshift variant we identified in *MSH6* is strikingly similar to other pathogenic variants known to cause Lynch Syndrome. This patient also has a personal history of precancerous colon polyps and multiple benign breast masses. Additionally, there is a family history that includes a sibling diagnosed with endometrial cancer. Unlike cHL, endometrial cancer has been strongly associated with Lynch Syndrome and variants in *MSH6*. Endometrial cancer has a prevalence of 3% in patients with Lynch syndrome and is the most frequently observed cancer in women harboring pathogenic *MSH6* variants, with a 41% risk of developing endometrial cancer by age 70^75,76^. Although cHL is not a cancer that is generally associated with Lynch syndrome, Wienand et al.^9^ also identified two hypermutated cases in a cohort of cHL patients. Wienand et al.^9^ indicate that the hypermutator phenotypes they observed were associated with mutation signatures that are consistent with microsatellite instability (COSMIC signatures 6 and 15), which is similar to our result. They also report somatic alterations in *MSH3*, *MSH2* and *ARID1A*, but do not report an analysis of germline variants. Our results suggest that hypermutation does occur in cHL patients, and this phenotype may be the result of germline cancer predisposition variants and could be important to guide therapy with immune checkpoint blockade.

In summary, we have shown that ultra-deep sequencing can be used to identify somatic variants in rare malignant HRS cells. We have further described the role that aberrant somatic hypermutation plays in cHL, and we have suggested a novel role of mutations in *IL4R* in the constitutive activation of STAT6. We also revealed that genes regulating Hippo signaling, the MAPK pathway, and the phosphoinositide signaling cascade may also play a role in cHL. We demonstrated the utility of ultra-deep sequencing in bulk cHL lymph node biopsies as an alternative to laborious cell isolation techniques. As sequencing costs continue to decrease, these methods have the potential to provide a platform to attempt larger cohort sequencing of primary cHL samples and open new avenues of research facilitating a practical approach to identify and correlate cHL mutations with clinical outcome.

## Supporting information

STROBE_check_list

supplemental methods and results

## Data Availability

The raw de-identified exome sequences described in this analysis will be deposited in the NCBI sequence read archive (SRA) via dbGaP under the accession phs001229

## Data Availability

The raw de-identified exome sequences described in this analysis will be deposited in the NCBI sequence read archive (SRA) via dbGaP under the accession phs001229. The amplicons used for the orthogonal validation experiment are included in the supplementary data files.

## Acknowledgements

The authors thank Alvin J. Siteman Cancer Center at Washington University School of Medicine and Barnes-Jewish Hospital and the Institute of Clinical and Translational Sciences (ICTS) at Washington University, for the use of the Tissue Procurement Core, which provided DNA isolation services. The Siteman Cancer Center is supported in part by an NCI Cancer Center Support Grant #P30 CA091842 and the ICTS is funded by the National Institutes of Health’s NCATS Clinical and Translational Science Award (CTSA) program grant #UL1 TR002345. The authors thank Grace Triska for her help with early clinical data organization. M.G. was supported by a National Institutes of Health, National Human Genome Research Institute grant (K99 HG007940). O.L.G. was supported by National Institutes of Health, National Cancer Institute grants (U01 CA209936, U24 CA237719, and K22 CA188163). This work was supported by the Siteman Investment Program Team Science Award (TAF, BK), the Larry and Winnie Chiang Lymphoma Fellowship (T.A.F., F.G.), the Paul Calabresi Career Development Award (K12 5K12CA167540-07; F.G.), the Siteman Cancer Center Research Development Award (T.A.F.), and the Foundation for Barnes-Jewish Hospital Steinback Fund (N.L.B.).

## Contributions

O.L.G., T.A.F., and M.G. conceptualized the study. M.G., O.L.G., and T.A.F. developed the experimental design with assistance from J.W., F.G., C.F., and R.S.F.. A.C., N.L.B., N.M-S., B.K., D.R-G, and T.A.F. acquired samples and clinical data. M.P.W., T.S., M.O’L., and J.W. performed laboratory experiments, sample organization, and/or clinical data organization. Y-S.L. and E.D. performed histological assessments. F.G., A.K., M.M., Z.S., E.D., A.S., F.M.R., C.A.M., L.T., K.K., C.R., J.W., H.A., A.K, M.G., and O.L.G. performed data analysis. F.G.,J.M., M.M.,T.A.F., M.G., O.L.G. prepared figures and tables. F.G., O.L.G., M.M., M.G. and T.A.F. wrote the manuscript with assistance from K.K.,D.R-G, N.M-S, and N.L.B. All authors approved the final version of the manuscript.

## Online Methods

### Patient sample acquisition, characteristics, and ethical considerations

All patients provided written informed consent for the use of their samples in sequencing as part of the Washington University School of Medicine (WUSM) Lymphoma Banking Program. The WUSM Institutional Review Board (IRB) approved protocols include: IRB 201108251, 201104048, 201110187. All human research activities are guided by the ethical principles in “The Belmont Report: Ethical Principles and Guidelines for the Protection of Human Subjects Research of the National Commission for the Protection of Human Subjects of Biomedical and Behavioral Research”.

For this project we included all fresh frozen excisional biopsies available in the bank from 2008-2015. Pathology review was performed on frozen lymph node samples to confirm the diagnosis of cHL (ED & YL). Nodular lymphocyte-predominant Hodgkin lymphoma (NLPHL) was not included. Non-malignant samples were collected (skin punch biopsies) and were included for germline analysis. Frozen sections (tumor and skin) were cut and used for genomic DNA isolation. The majority of samples included in this study were untreated at the time of biopsy. In these cases, the sample included was the diagnostic excisional biopsy or core needle biopsy (27 untreated cases). There were also 4 relapses included. In two cases the second relapse biopsy was sequenced and for the remaining two cases, the first and fourth relapse biopsy was sequenced. Basic demographics and clinical features are described in Table 1.

### DNA isolation, library preparation and sequencing

DNA was isolated using a Gentra Puragene kit at the Washington University Tissue Procurement Core facility. Automated dual indexed libraries were constructed with 30-250ng of genomic DNA utilizing the KAPA HTP Library Kit (KAPA Biosystems) on the SciClone NGS instrument (Perkin Elmer) targeting 250bp inserts. Three libraries were constructed per sample, each with a unique index. Libraries from the same patient (3 from the normal and 3 from the tumor) were pooled prior to hybridization, yielding a 3µg library pool. Each library pool was hybridized with the xGen Exome Research Panel v1.0 reagent (IDT Technologies) which spans a 39 Mb target region (19,396 genes) of the human genome. The concentration of each captured library pool was accurately determined through qPCR (Kapa Biosystems) to produce cluster counts appropriate for the HiSeq X platform (Illumina). 2x151bp paired end sequence data were generated with a target of approximately 100Gb per sample and target mean coverage of approximately 1000x.

### Sequence alignment, variant calling and filtering

Sequence analysis and data management was performed using the Genome Modeling System (GMS)^1^. Briefly, paired-end reads were aligned to human reference sequence GRCh38 using BWA-MEM v0.7.10-r789^2^ via the Speedseq analysis platform (v0.10)^3^. Duplicate marking was completed using samblaster v0.1.22^4^ and alignments were sorted using sambamba v0.5.4^5^. Somatic variants were called using SAMtools^6^, SomaticSniper^7^, VarScan^8^, MuTect^9^, Strelka^10^, Pindel^11^, and GATK^12^.

Following variant calling, all SNVs and INDELs were annotated using an in house annotation pipeline^1^. After variant annotation, all variants were filtered to remove common variants, sequencing errors, and pipeline artifacts. We required all variants to have a minimum of 50x depth in both the tumor and normal samples. We excluded sites with the following characteristics: normal VAF >= 5%, tumor VAF <= 0.5% and sites with <= 5 variant supporting reads in the tumor. We filtered sites based on their predicted consequences. We removed sites from the 5’ and 3’ UTRs, intronic sites, synonymous variants, and sites annotated to non-coding transcripts. To remove artifacts and sequencing errors generated in this dataset by the pipelines we required all variants to be called in at least 2 of the 3 libraries for each sample. Further, within each library a variant was required to have at least 3 reads of support and a VAF >0.5%. As a further attempt to remove pipeline artifacts and sequencing errors, all variants were filtered against the 31 normal samples from this study. In this step, SNVs were removed if they had >4 variant reads and >1% VAF in 2 or more normal samples and INDELs removed if they had >3 variant reads and >1% VAF in 3 or more normal samples. Variants were removed if their global minor allele frequency was >0.001 ExAC release 0.2.^13^

Following all automated filters, experienced analysts reviewed all variants in IGV^14^ using standard operating procedures^15^ to identify sequencing errors, variant caller artifacts, false positives (e.g., germline variants) and other systematic anomalies.

### HaloPlex Validation

Following variant calling, variant automated filtering and manual review, we designed a custom capture reagent using the HaloPlex HS Target Enrichment System (Agilent Technologies) to validate the somatic variants. We included 1,842 out of 4,692 SNVs and INDELs that passed manual review. The variants that were included on the HaloPlex panel were all passing sites from all samples except where amplicons could not be designed (i.e. mitochondrial genes and repetitive regions), and a subset of sites from one sample (HL-513) with a very large number of variants. From this hypermutator, 834 out of 3,684 sites were included on the HaloPlex panel. We additionally tiled across 30 genes selected from the most recurrent genes in the cohort and genes known to be recurrently mutated in Hodgkin lymphoma (Supplementary Table 5).^16,17^ The HaloPlex reagent was designed using the Agilent SureDesign platform (Supplementary File 1). All probes were designed with two indices, a unique molecular barcode (UMI) to allow for error-corrected sequencing, and a sample index to allow for sample multiplexing (sample index).

HaloPlex libraries were created, sequenced, and processed using methods similar to previous reports,^18,19^ and the HaloPlex HS Target Enrichment System manufacturer protocol (Agilent Technologies, CA). Up to 500ng of genomic DNA was first digested using a mixture of restriction endonucleases in the HaloPlex kit. Library quality was assessed with the Agilent 2100 Bioanalyzer. Fragmented genomic DNA was then hybridized to the HaloPlex HS probe library. Hybridized genomic DNA fragments were ligated to close nicks in the probe-target DNA hybrids, captured with streptavidin, and amplified with PCR (22 cycles), creating read families, each with its own unique molecular barcode index. Library concentration was assessed with qPCR according to the manufacturer’s protocol (Kapa Biosystems). Both tumor and normal samples were interrogated in the HaloPlex validation experiment.

Libraries were normalized, pooled, and sequenced on the HiSeq 4000. Eight samples were sequenced with 2 samples/lane and the remaining samples were sequenced with 3 samples/lane. HaloPlex sequence data was processed similar to the methods described previously.^18^ Barcoded FASTQ data was demultiplexed and then reads were trimmed using Flexbar.^20^ Trimming was performed to remove systematic errors introduced to the end of reads by HaloPlex chemistry. Reads were then aligned with BWA MEM v0.7.10-r789.^2^

All SNVs were evaluated using BarCrawler, a custom GATK-based tool. (https://github.com/abelhj/gatk/tree/master/public/external-example/src/main/java/org/abelhj). As described in Wong et al,^18^ background noise calculation was performed on a position-by-position basis for each identified SNV. At each site read counts were gathered from all other samples. A Fisher’s exact test, implemented in the R statistical environment, was used to compare the reference and variant read counts at a site to the number of reference and variant reads at that site in all other samples. The *p*-value for this test was retained. Multiple testing correction was then applied with the *p*.adjust function (base R; default parameters - “holm”^21^). Those variants with an adjusted *p*-value of less than 0.1 were retained. The same process was then repeated with subsequent background calculations excluding all variants retained in previous rounds until no new variants were identified.

Following the final iteration of the background noise correction, SNVs with an adjusted p-value of 0.05 were carried forward. Several other parameters were considered to validate a SNV. SNVs were required to have an error corrected depth >= 100 reads in the tumor and the normal, and an adjusted p-value >0.05 in the normal. In some instances the normal sample did not have sufficient HaloPlex data to evaluate whether or not a site was present in the normal. In these cases we designated the site as a ‘tumor-only’ validated site if sufficient tumor depth (100x) was reached and a tumor p-value <= 0.05 was observed. Finally, as stated above only 834 of the 3,684 sites from the hypermutated patient were included on the HaloPlex reagent. We determined that only 12 of the 834 interrogated sites failed validation. Because this sample had a validation rate ∼99% and few failing sites upon which a custom validation model could be trained, we simply considered the additional 2,850 SNVs from this patient as high confidence sites and were included in all subsequent analyses.

Indels were assessed using “consensus bams” - alignments created based on error corrected HaloPlex data. Consensus reads were generated from demuxed sequencing data via a wdl workflow that uses WalkerTRConsensus_wk5 (https://github.com/abelhj/gatk/blob/master/public/external-example/target/external-example-1.0-SNAPSHOT.jar).

Briefly, reads were trimmed of adapter sequences via cutadapt (-m 30 -u 3) and aligned with BWA-MEM. WalterTRConsensus_wk5 was then applied (-dcov 1000000 -maxNM5 -mmq 10) to generate consensus reads. Read families with less than 3 individual reads with the same UMI were removed and not used to create consensus reads. All INDELs were manually inspected using IGV^15^ and bam-readcount (<u>genome/bam-readcount: count DNA sequence reads in BAM files</u>) was used to assess the number of variant supporting reads and the VAF for the variant. For INDELs we required: a consensus bam tumor and normal depth > 5 consensus reads; > 1 consensus tumor read of support; < 20 consensus normal bam reads of support; consensus tumor VAF > 0.01%; a consensus normal VAF < 5%. Similar to the SNVs, if the normal consensus bam could not be evaluated, we required sufficient tumor bam depth (5 consensus reads), 1 variant read of support in the tumor and a tumor VAF > 0.01%.

### De novo variant calling

Using the consensus bams described above we attempted to call variants within the entire HaloPlex analysis space. To accomplish *de novo* variant calling we created a custom low VAF variant calling pipeline by modifying an existing GMS^1^ variant calling workflow. Briefly, all post processing filters were removed, leaving only the filtering that occurs at the variant caller level using the default settings (https://github.com/fgomez02/analysis-workflows/blob/9c9e6a6a48eb321804ce772a2c2c12b4f2f32529/definitions/pipelines/detect_variants.cwl).

After variants were called, all de novo variants were filtered for basic variant quality. Variants with less than 5 consensus reads in the tumor and normal were removed. Sites were required to have a normal VAF < 5%, and a tumor VAF > 0.5%. We also required < 5 variant supporting reads (consensus reads) in the normal and >= 2 variant supporting reads in the tumor. All *de novo* variants were annotated using the Ensembl Variant Effect Predictor (VEP) tool (Ensembl v93). Then, we filtered these variants by consequence using similar consequence filters described for the exome filtering pipeline. All variants that were previously known to us were removed. We also intersected all remaining variants with the HaloPlex analysis space using bedtools,^22^ keeping only variants in regions where probes were designed. Following automated filters all remaining variants were manually reviewed in IGV^14,15^. During manual review the consensus tumor and normal bam files were loaded, as well as the exome tumor and bam file. Variants were passed if support was seen in the tumor consensus bam as well as the tumor exome bam file. Because we called and annotated new variants in the *de novo* variant calling exercise, we updated the consequence annotations of all exome variants using Ensembl Variant Effect Predictor (VEP) tool (Ensembl v93) for all sites that passed validation so that all sites would be annotated consistently. Additionally, it should be noted that 7 variants in *TCIRGI*, *CEP290*, *SMAD3*, *CEP131*, *COMP*, *LMTK3*, and *DOCK3*, that originally failed the orthogonal validation were called again in the *de novo* variant calling exercise. After manual review of all available data these sites were rescued and included in the final analyses.

### Hotspot variant read counting

To interrogate our data for known hotspots in cHL, we used bam-readcount (https://github.com/genome/bam-readcount) on 10 known variants (6 variants in *STAT6*, 3 variants in *XPO1* and 1 variant in *NFKBIE* - Supplementary Table 1) in all consensus tumor and normal bams. Sites were required to have a tumor VAF >= 0.5%, normal VAF < 5%, a tumor var count > 5 reads, and a normal var count <= 2 reads. We also required a total of 5 reads in the tumor and normal. Additionally, similar to the consensus *de novo* variant calling, all variants that passed the automated filters were manually reviewed. All variants at hotspot locations previously unknown to us, that passed all filters, were included.

### Variant Analyses

#### Significantly mutated gene analysis

We identified significantly mutated genes (SMGs) using MuSiC.^23^ The region of interest was restricted to the coding space covered by the exome reagent. Additionally, two bases were added to the beginning and end of each exon to account for splice sites. All default parameters were used. Significance was determined by comparing the minimum FDR value from the convolution and likelihood ratio tests to our predetermined threshold - 0.05.

#### Pathway analysis

To identify pathways that are impacted by our somatic mutations we first downloaded the C2 curated gene set collection from MSigDB^24,25^. This gene set includes canonical pathways from several online databases including BioCarta, KEGG, and Reactome. This gene set also includes genes from the biomedical literature that have been shown to be related to important biological and clinical states (e.g. cancer metastasis or drug resistance). We also included a list of 1,600 known human transcription factors^26^, a list of known regions of off target AID activity in DLBCL and FL,^27,28^ and selected WikiPathways^29^ (e.g. Hippo regulating pathways). Note that the MAPK signaling pathway was defined as the genes included in the KEGG MAPK signaling pathway with the addition of the following genes: *PCDH7*^30^*, TRAF7*^31^, *ARAF, DAB2IP, SHC3, IRS2, KSR2,* and *KSR1*^32^. We identified the pathways from these gene sets that were impacted by our most recurrently mutated genes by searching for genes mutated in 3 or more patients. When a recurrently mutated gene was mapped to more than one pathway, manual inspection was used to retain the pathway(s) most likely to be relevant to cHL. Following the creation of our “pathways of interest” data set, we further annotated all genes mutated in all patients to these pathways.

#### Activation Induced Deaminase Analysis

In the genes we identified as potential targets of aberrant AID activity in the pathway analysis described above ^27,28^ (Supplementary Table 4) we tested whether these loci contain somatic mutations (SNVs) that are likely to be the product of aberrant AID activity. SNVs were considered products of off target AID activity if they met the following criteria: the SNV must have a <u>C</u> (or G) as the reference allele, be within 2 Kb of transcript start sites (TSS), and be within the WR<u>C/G</u>YW AID sequence motif. To identify whether a mutation is within an AID sequence motif we used custom software (Fasta-Region-Inspector(FRI): https://github.com/Matthew-Mosior/Fasta-Region-Inspector).

To assess the significance of the number of AID target mutations we identified, we used additional custom software (Fasta-Region-Randomizer: https://github.com/Matthew-Mosior/Fasta-Region-Randomizer) to create 100,000 simulated datasets that contain the same number of SNVs observed within each target gene. In these simulations, the genomic space that was considered included all exons of each gene (i.e. the exonic space included in the exome capture). We then determined the observed number of putative AID mutations in each simulated dataset using FRI. To assess whether the overall number of off-target AID mutations is significantly different from random expectations we calculated how many times our total number of actual AID mutations (or greater) was observed in the simulated data.

#### Mutation signature analysis

Mutation signatures were assigned using the deconstructSigs R package.^33^ The reference signatures evaluated were the 30 COSMIC v2 signatures of mutational processes in human cancers (http://cancer.sanger.ac.uk/cosmic/signatures). Samples that contained >= 50 SVNs were included in this analysis.

#### Germline Analysis

Sequencing data from the hypermutated patient was interrogated for germline variants within genes that are involved in mismatch repair and base excision repair (Supplementary Table 2). Sequence data was aligned using the same methods previously described for the WashU post harmonization alignment and data processing pipeline.^34^ Briefly, reads were aligned using BWA MEM v0.7.15-r1140^2^, MC (mate CIGAR) and MQ (mate mapping quality) tags were added using samblaster v0.1.24^4^, alignments were sorted using sambamba^5^, and duplicates were marked using Picard (http://broadinstitute.github.io/picard/). Variants were called using the GATK Haplotype caller (v3.5.0) and the GATK Genotype Caller v3.4-0-gf196186.^35^

Following variant identification, variants were annotated using VEP (Ensembl v93). To create a high quality variant list, variants were required to have a depth of coverage ≧20 reads, ≧ 5 variant supporting reads, and a VAF ≧ 2.5%. Additionally, to remove variants unlikely to impact gene function we removed variants annotated as located in the 5’ or 3’ UTR, synonymous, non-coding, downstream, nonsense mediated decay variants, and variants located in gene regions that have been annotated as regulatory regions. We also applied a gnomad (v2.1)^36^ population allele frequency where we removed variants with a MAX_AF > 0.1% filter. Following all filters the remaining variants were further examined through manual review^14^.

