## supplemental methods and results for "Ultra-Deep Sequencing Reveals the Mutational Landscape of Classical Hodgkin Lymphoma"

### Supplementary Information for Ultra Deep Sequencing Reveals the Mutational Landscape of Classical Hodgkin Lymphoma

#### Supplementary Results

*Clinical Associations and EBV association tests*

Clinical history and/or outcome data were available for all 31 patients. We evaluated the difference in progression free survival (PFS) in patients stratified by mutation status at genes mutated in 3 or more patients. We did not observe an association between PFS and mutation status with a p value < 0.1. The top two associations we observed were at *LMTK3* (p=0.11) and *CDH5* (p= 0.17) (Supplementary Table 3).

Our methods for determining EBV status were mostly concordant; 5/5 EBER positive samples appeared positive in the competitive alignments. Among the EBER negative patients, 17/21 were also observed as negative using the alignment method. Four patients were determined negative by EBER but appeared positive using DNA alignments. There were no patients that appeared negative in the DNA alignments but were determined to be positive using EBER ISH. We did not observe a significant difference between mutation burden and EBV status determined by EBER (p=0.11) or competitive alignment (p= 0.82; Supplementary Figure 8).

A correlation was not observed between median mutation VAF per sample and the approximate total number or RS cells. However, it should be noted that the sequenced frozen tissue was not amenable to rigorous histologic estimates of RS cell numbers, so our estimate of RS cell count is unlikely to fully represent the number of sequenced RS cells.

#### Supplementary Methods

*Detection of EBV and association of EBV status with mutation burden*

In-situ hybridization for Epstein-Barr virus (EBV) mRNA (EBER) was used to detect the presence of Esptien Barr virus within each lymph node biopsy. Kallisto[1] was used to determine the presence of EBV DNA by performing competitive pseudo alignments to the human reference genome (GRCh38) and multiple viral genomes (EBV, human papillomavirus, and hepatitis B). To verify that our pseudo aligned reads were correctly aligned to EBV, we used BLASTn to search for all matching reads and verified that the EBV reference was among the highest concordances. Finally, to confirm the presence of EBV viral DNA all samples were competitively aligned to the human and EBV reference genomes using BWA mem[2]. These alignments were then visualized in IGV[3] where the presence of EBV reads was confirmed.

We used a two tailed T-test to test whether the mutation burden (i.e. average number of mutations) across all samples that are EBV positive is significantly different from the mutation burden in EBV negative cases. This test was performed using EBV status determined via EBER in situ hybridization and EBV status determined using the competitive alignment methods described above.

*Survival Analysis*

For survival analysis, patients were stratified by mutation status (mutated vs. wild-type) for all genes mutated in 3 or more patients. Time-to-event analyses were performed using a log-rank test to identify significant PFS (progression-free survival) differences between patients with mutations in a particular gene and patients without mutations.

#### Supplemental Figures

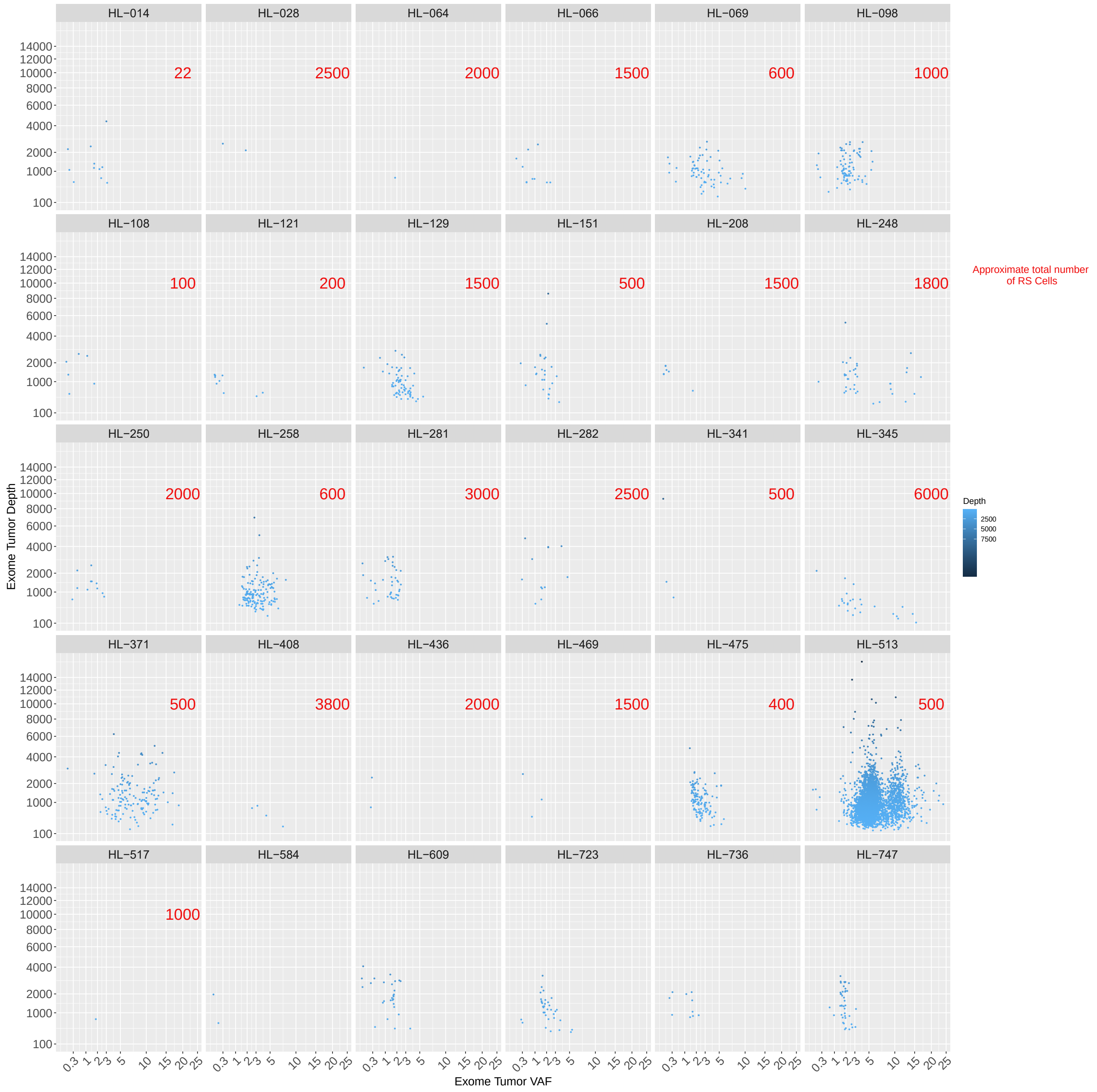


*Supplementary Figure 1. Exome VAF and Depth of Coverage*

A summary of all variants presented, plotted by sample. Each dot represents a variant. The variant allele frequency (VAF) and depth of coverage is shown. Dots are shaded based on exome depth. The numbers in red are the approximate number of Reed Sternberg cells (RS). We were unable to determine an approximate RS count for 5 samples. Patient HL-157 is not included because mutations from this patient were not included in any analyses.

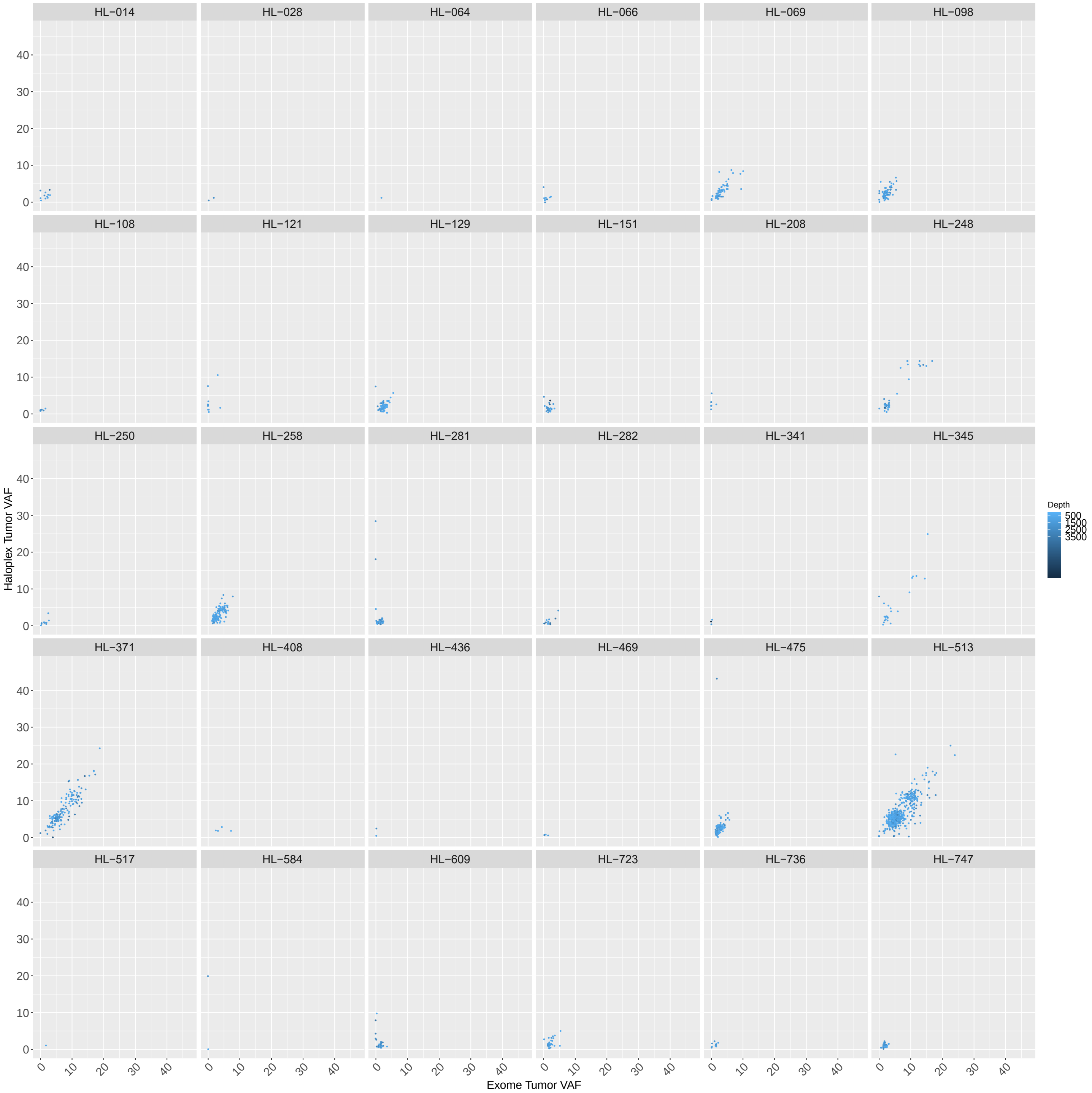


*Supplementary Figure 2. Exome VAF and HaloPlex Tumor VAF*

A summary of the HaloPlex and exome variant allele frequencies (VAF) for all variants presented here, plotted by sample. Each dot represents a variant. Dots are shaded based on exome depth. Patient HL-157 is not included because mutations from this patient were not included in any analyses

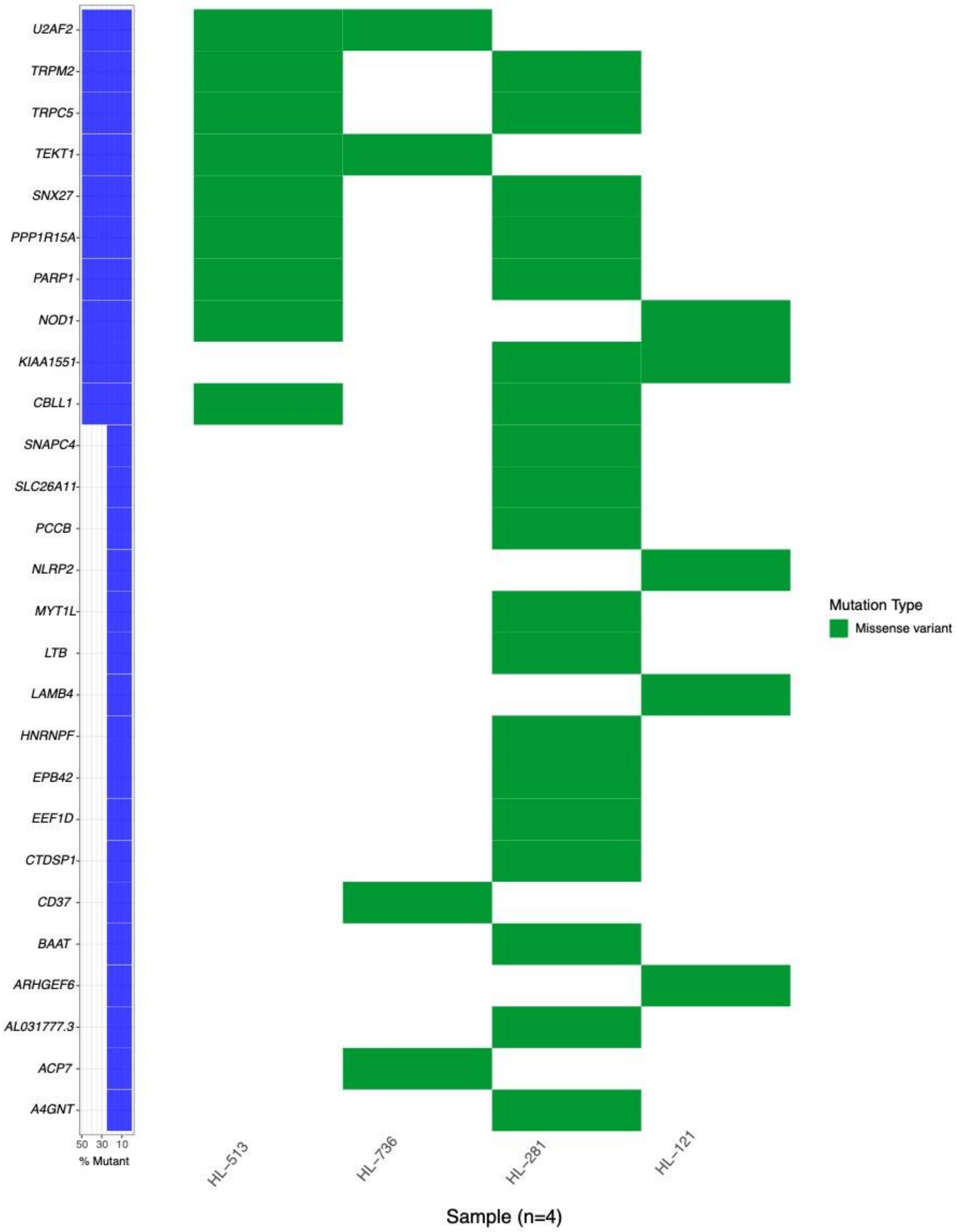

U2AF2
TRPM2
TRPC5
TEKT1
SNX27
PPP1R15A
PARP1
NOD1
KIAA1551
CBLL1
SNAPC4
SLC26A11
PCCB
NLRP2
MYT1L
LTB
LAMB4
HNRNPF
EPB42
EEF1D
CTDSP1
CD37
BAAT
ARHGEF6
AL031777.3
ACP7
A4GNT
50 30 10
% Mutant
HL−513
HL−736
HL−281
HL−121
Sample (n=4)
Mutation Type
Missense variant

*Supplementary Figure 3. Summary of Recurrently Mutated Genes for Relapsed Samples*

The frequency and type of mutations found in the non-hypermutated relapse samples. Also included are genes mutated in the hypermutated sample and one additional relapse sample. Each column represents a relapse patient. The bar graph on the left summarizes the frequency of mutations for that gene across the relapse samples.

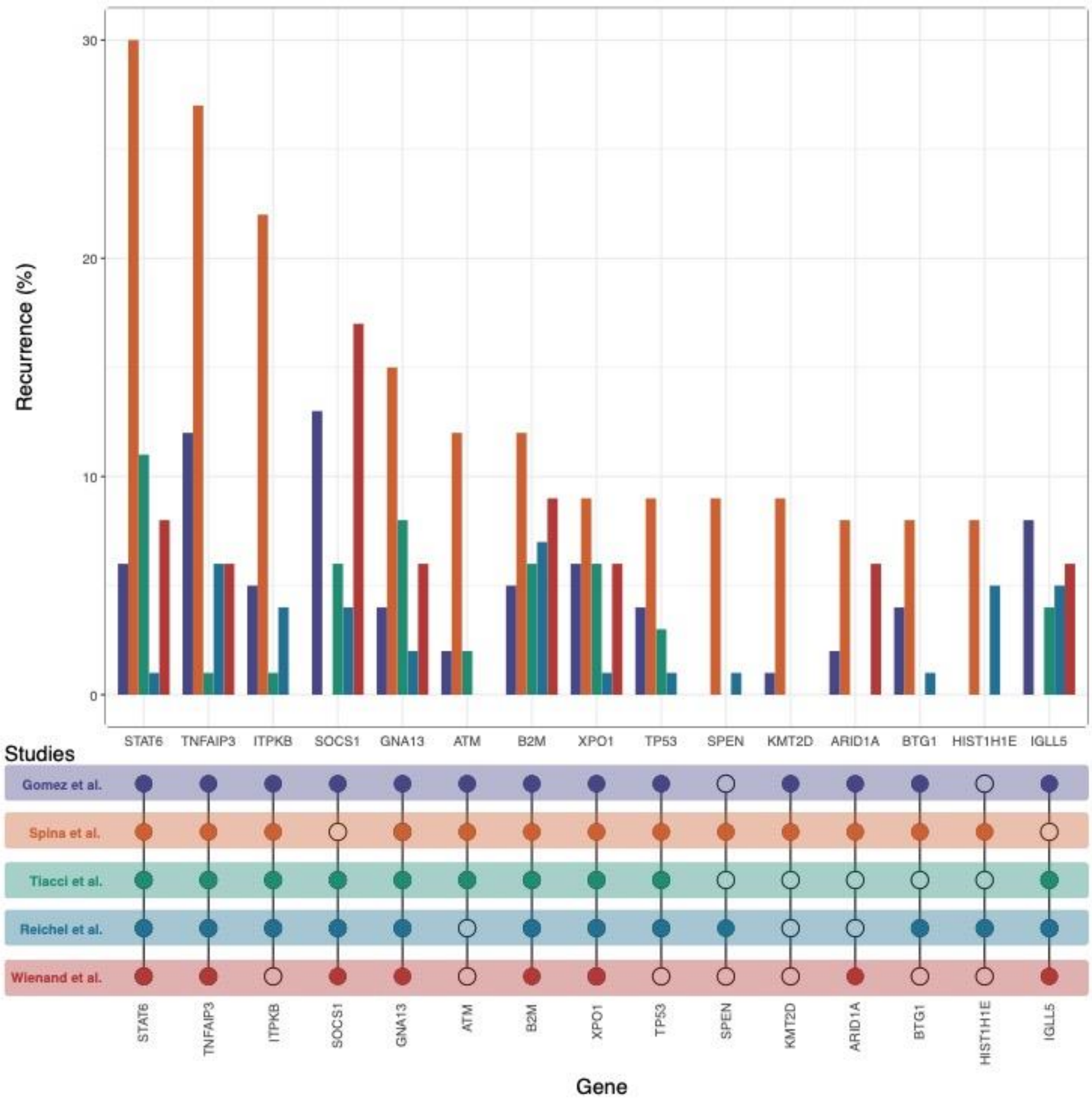

*Supplementary Figure 4. Comparison of Genes Shown to be Mutated in Studies of Adult cHL*

The genes summarized here are the 15 most recurrently mutated genes across the 5 studies of the genomic landscape of adult cHL.[4–7] Each bar represents the recurrence of a particular gene, colored by study. The lower plot indicates whether each included study reported mutations in the selected genes.

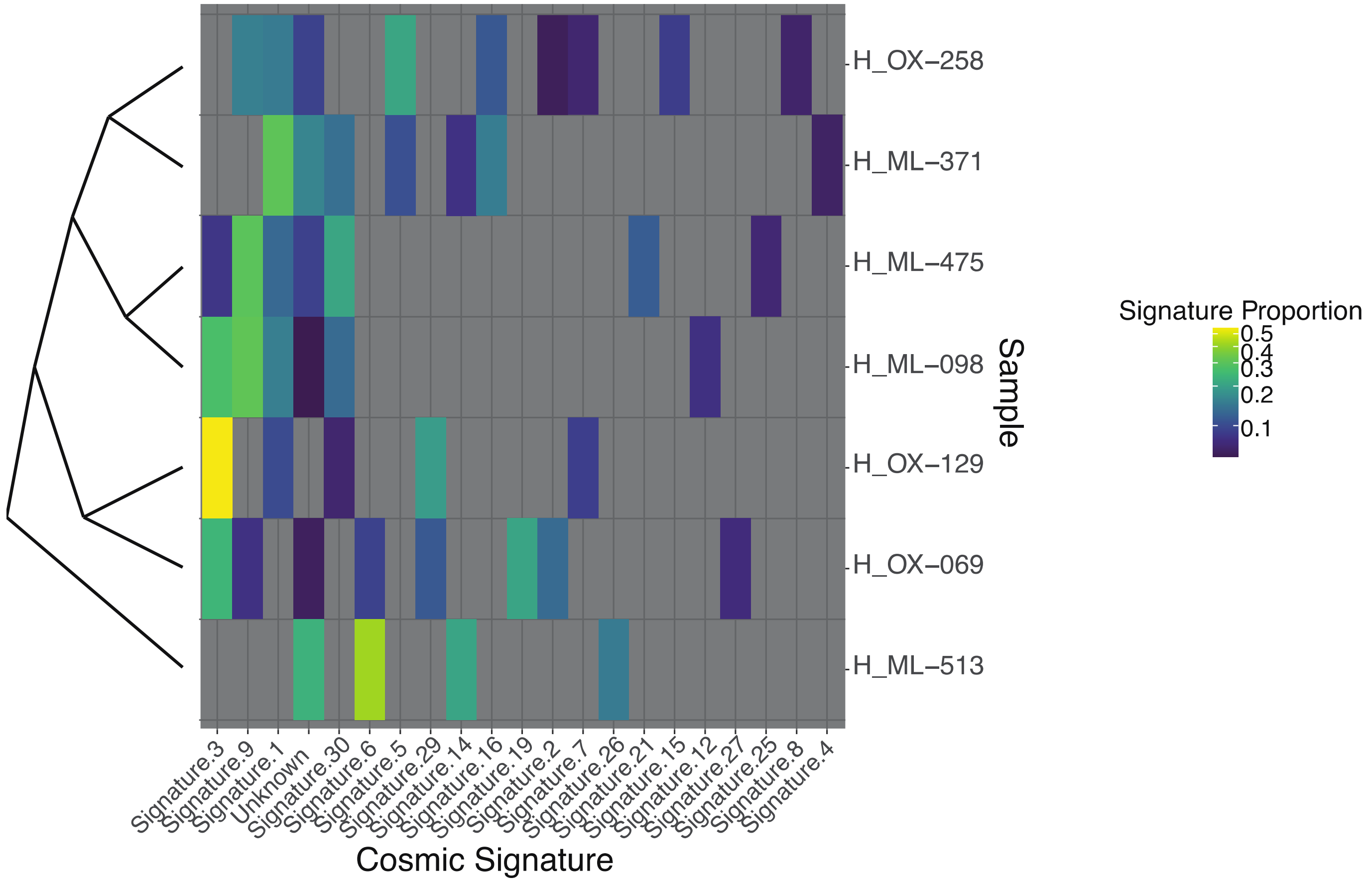


*Supplementary Figure 5. Observed COSMIC v.2 mutation signatures*

Patients included in this analysis had at least 50 somatic mutations. Shading represents the observed proportion of a particular signature out of all signatures observed in that sample. Dendrogram represents sample relatedness based on similarity of mutation signatures.

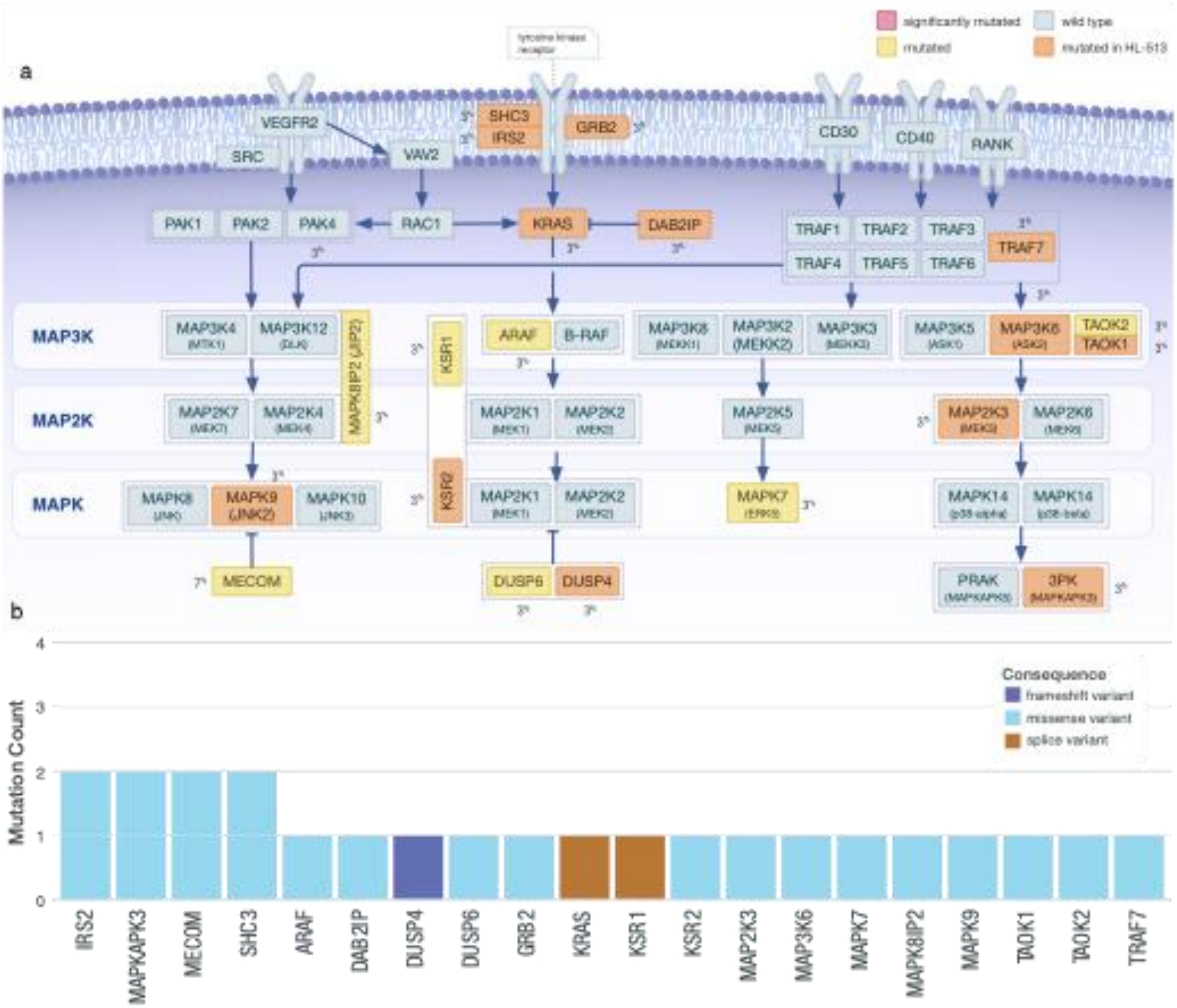

a
b
significantly mutated
wild type
mutated
mutated in HL-513
tyrosine kinase receptor
VEGFR2
SRC
VAV2
SHC3
IRS2
GRB2
CD30
CD40
RANK
PAK1
PAK2
PAK4
RAC1
KRAS
DAB2IP
TRAF1
TRAF2
TRAF3
TRAF4
TRAF5
TRAF6
TRAF7
MAP3K
MAP2K
MAPK
MAP3K4
MAP3K12
MAPK8IP2 (JIP2)
KSR1
KSR2
ARAF
B-RAF
MAP3K8
MAP3K2 (MEKK2)
MAP3K3
MAP3K5
MAP3K6
TAOK2
TAOK1
MAP2K7
MAP2K4
MAP2K1
MAP2K2
MAP2K5
MAP2K3
MAP2K6
MAPK8
MAPK9 (JNK2)
MAPK10
MAPK7
MAPK14
MECOM
DUSP6
DUSP4
PRAK (MAPKAPK5)
3PK (MAPKAPK3)
Consequence
frameshift variant
missense variant
splice variant
Mutation Count
0
1
2
3
4
IRS2
MAPKAPK3
MECOM
SHC3
ARAF
DAB2IP
DUSP4
DUSP6
GRB2
KRAS
KSR1
KSR2
MAP2K3
MAP3K6
MAPK7
MAPK8IP2
MAPK9
TAOK1
TAOK2
TRAF7

*Supplementary Figure 6. Diagram of observed mutated components of the MAPK signaling cascade*

a) Genes mutated only in the hypermutated sample are shown in orange; genes mutated in at least one non-hypermutated sample are shown in yellow; genes that were identified as SMGs are shown in red. The frequency of the gene mutated across the cohort is shown as a percent. b) The total number and type of mutations observed are shown in inset bar chart.

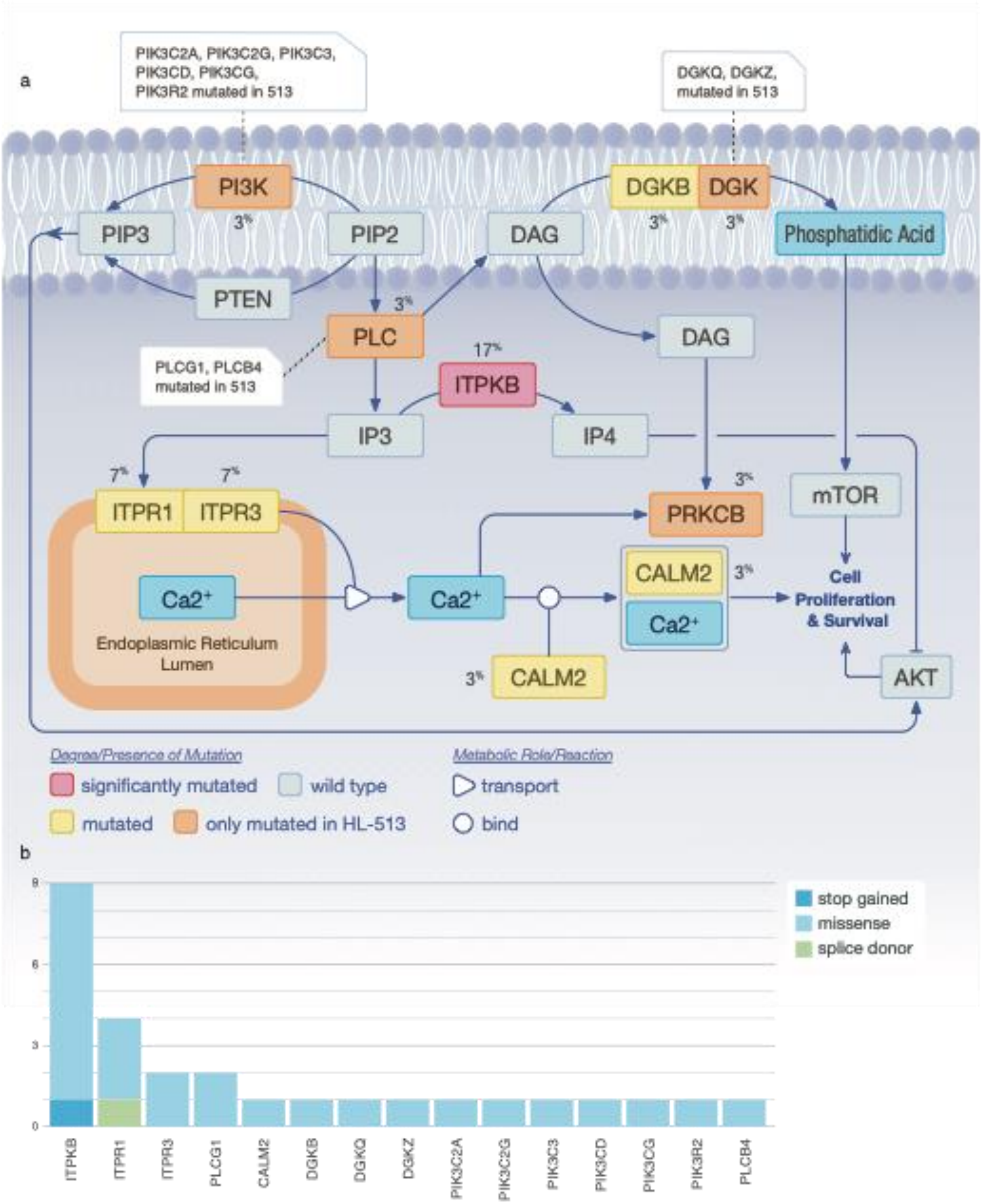

a
PIK3C2A, PIK3C2G, PIK3C3, PIK3CD, PIK3CG, PIK3R2 mutated in 513
DGKQ, DGKZ, mutated in 513
PI3K
PIP3
PIP2
PTEN
DAG
DGKB
DGK
Phosphatidic Acid
PLC
PLCG1, PLCB4 mutated in 513
ITPKB
IP3
IP4
DAG
ITPR1
ITPR3
PRKCB
mTOR
Ca2+
Endoplasmic Reticulum Lumen
Ca2+
CALM2
Ca2+
Cell Proliferation & Survival
CALM2
AKT
Degree/Presence of Mutation
significantly mutated
wild type
mutated
only mutated in HL-513
Metabolic Role/Reaction
transport
bind
b
stop gained
missense
splice donor
ITPKB
ITPR1
ITPR3
PLCG1
CALM2
DGKB
DGKQ
DGKZ
PIK3C2A
PIK3C2G
PIK3C3
PIK3CD
PIK3CG
PIK3R2
PLCB4

*Supplementary Figure 7.  Diagram of observed mutated components of the PI3K signaling cascade*

a) Genes mutated only in the hypermutated sample are shown in orange; genes mutated in at least one non-hypermutated sample are shown in yellow; genes that were identified as SMGs are shown in red. The frequency of the gene mutated across the cohort is shown as a percent. b)The total number and type of mutations observed are shown in inset bar chart.

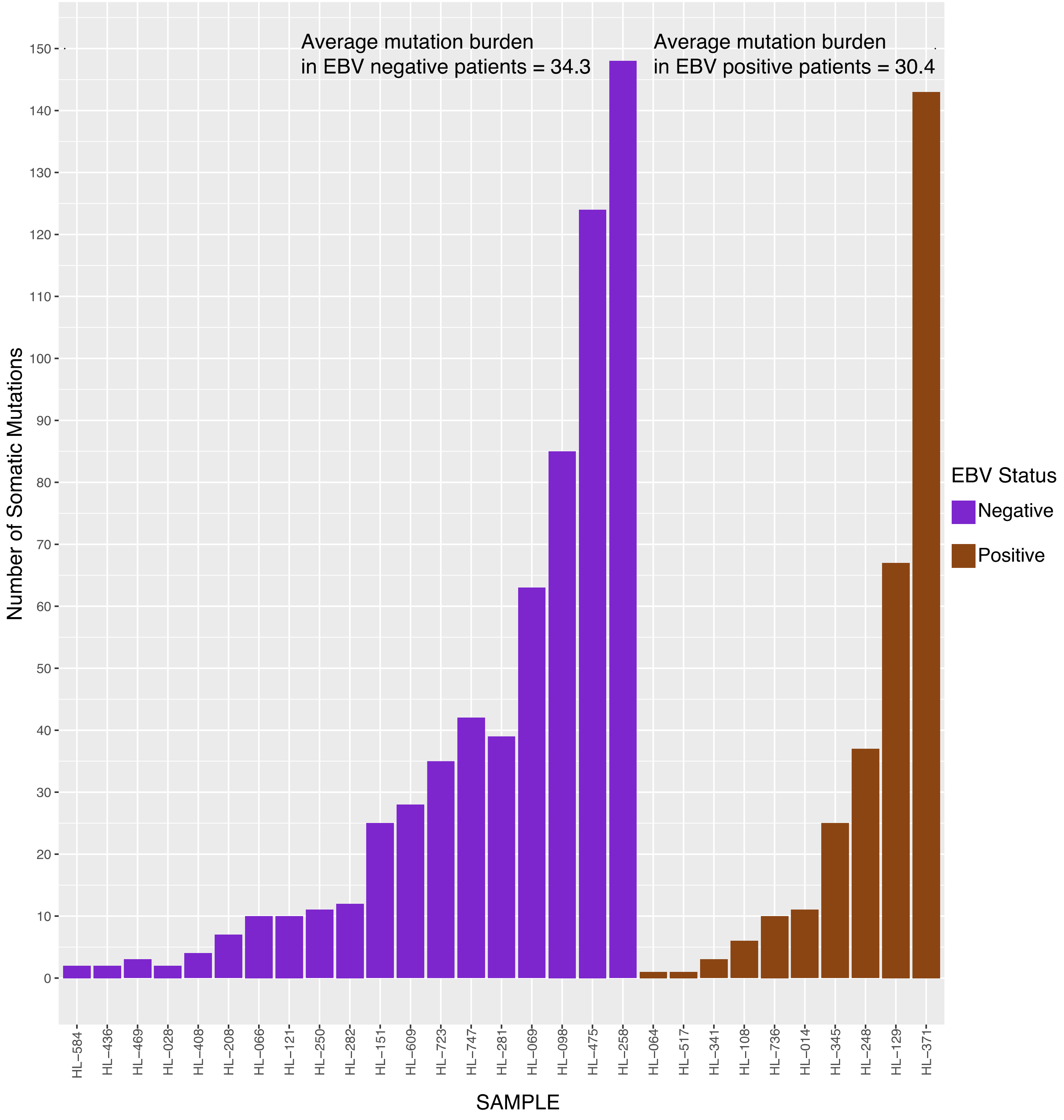

*Supplemental Figure 8. Comparison of mutation burden and EVB status*

Mutation burden (mutation count) in samples that were found to be EBV positive (purple) or EBV negative (brown) using competitive alignment (Methods). The competitive alignment was largely concordant with EBV status determined using EBER ISH. Comparison of mutation burden between EBV positive and EBV negative patients using a t-test , excluding HL-513, indicates that there is not a significant difference in the mutation burden between the two groups (p=0.82).
